# The antibody landscapes against Group 1 and 2 influenza virus hemagglutinin following AS03 and MF59 adjuvanted H5N1 vaccination

**DOI:** 10.1101/2021.07.09.21259780

**Authors:** Johannes B. Goll, Aarti Jain, Travis L. Jensen, Rafael Assis, Rie Nakajima, Algis Jasinskas, Lynda Coughlan, Sami R. Cherikh, Casey E. Gelber, D. Huw Davies, Philip Meade, Daniel Stadlbauer, Shirin Strohmeier, Florian Krammer, Wilbur H. Chen, Philip L. Felgner

## Abstract

Current seasonal and pre-pandemic influenza vaccines induce short-lived predominantly strain-specific and limited heterosubtypic responses. To better understand how vaccine adjuvants AS03 and MF59 may provide improved antibody responses to vaccination, we interrogated serum from subjects who received 2 doses of inactivated monovalent influenza A/Indonesia/05/2005 vaccine with or without AS03 or MF59 using hemagglutinin (HA) microarrays. The arrays were designed to reflect both full length and globular head HA proteins derived from 17 influenza A subtypes (H1 to H16 and H18) and influenza B strains. We observed significantly increased strain-specific and broad homo- and hetero-subtypic antibody responses with both AS03 and MF59 adjuvanted vaccination with AS03 achieving a higher titer and breadth of IgG responses relative to MF59. Adjuvanted vaccine was also associated with the elicitation of stalk directed antibody. Finally, we established good correlation of the array antibody responses to H5 antigens with standard hemagglutination inhibition and microneutralization titers.

## INTRODUCTION

Avian influenza viruses represent a continuous pandemic threat, as illustrated by the frequent emergence of novel reassortant viruses which have resulted in sporadic spillover events from the zoonotic reservoir in the past few decades.^1, 2, 3, 4, 5^ These antigenically distinct influenza viruses are categorized into subtypes (i.e., H5, H7) based on phylogenetic characterization and sequence homology of the hemagglutinin (HA) gene, and are further sub-divided into clades. A key component of U.S. pandemic preparedness has been ongoing surveillance of prominent influenza viral clades, which may lead to their selection for development into vaccines which can be added to the National Pre-Pandemic Influenza Vaccine Stockpile (NPIVS).^6^ The rationale behind the stockpile is that a vaccine from pre-pandemic subtype viruses can provide partial cross-protection,^7^ thereby benefiting vaccinated priority groups before a better-matched vaccine against the pandemic strain becomes available. The NPIVS program currently contains multiple pre-pandemic influenza antigens, representing various H5Nx and H7N9 avian influenza viruses.

Inactivated subvirion influenza vaccines against avian HA strains have demonstrated poor immunogenicity in unprimed populations.^8^ Therefore, the NPIVS program also maintains two immune-stimulating adjuvants (AS03 and MF59). These adjuvants are intended to be deployed with a respective vaccine antigen in a mix and match strategy to provide better immune responses to vaccination, which might translate to dose-sparing of the limited supply and faster onset, greater breadth, and/or longer duration of protection with vaccination.^9, 10, 11^ One of the present gaps in knowledge is an understanding of the effect of these adjuvants on the breadth of the antibody responses elicited by pre-pandemic vaccines.

Influenza HA is a major target for humoral immune responses, with antibodies largely directed towards the immunodominant, but antigenically variable head domain (HA1). In contrast, the HA stalk domain (HA2) is highly conserved, but is immunosubdominant.^12, 13^ The standard approach to the evaluation of the antibody-mediated immune responses to influenza vaccination has been the measurement of antibodies by the hemagglutination inhibition (HAI) assay. Because there is no correlate of protection for avian influenza viruses, a microneutralization (MN) assay can provide an assessment of the functional antibodies elicited. Both the HAI and MN assays measure antibodies to specific viral strains, such that multiple independent HAI and MN assays need to be performed for each heterologous drifted viral strain of interest.

In recent years, it has been determined that non-neutralizing, HA stalk-specific antibodies, which display breadth of reactivity by ELISA, can confer protection from heterosubtypic influenza virus challenges in animal models.^14, 15, 16, 17^ Furthermore, such broadly reactive antibodies have recently been proposed as a correlate of protection in human cohort studies^18^ of natural influenza virus infection. Therefore, there is growing interest in identifying adjuvants that are capable of inducing such broadly reactive stalk-specific immune responses with the potential to confer breadth of protection against diverse influenza viruses.^19^ One novel method for more comprehensively assessing the breadth of antibody responses are protein microarrays. We constructed two sets of influenza-specific high-density protein microarrays which comprised purified HA proteins derived from 17 influenza A virus subtypes (H1 to H16 and H18) and influenza B virus strains. The first array was constructed to study the breadth of responses by including all available HA subtypes. The second array was similarly assembled, but used more conformationally correct stabilized H5 trimers, when available. Our present study was designed to measure the landscape of the antibody responses to vaccination using the protein microarray approach in response to vaccination with an inactivated, monovalent, subvirion influenza A/Indonesia/05/2005 (H5N1) strain vaccine when administered alone (unadjuvanted) or with AS03 or MF59 adjuvant (three possible vaccine groups). Importantly, using stabilized trimeric headless stalk protein (HA2) in competitive inhibition assays, we indirectly assessed the elicitation of stalk-directed antibodies in response to vaccination with or without AS03 or MF59 adjuvant. Finally, the concordance between microarray subtype specific antibody levels and HAI and MN titers were evaluated.

## RESULTS

### Experimental design

An overview of the experimental design is provided in Figure 1. The analysis cohort consisted of approximately 2/3 males and 1/3 females (Supplemental Table 1A) with an age range of 19 to 47 years and a median age of 27-28 years (Supplemental Table 1B). The concurrent sera from day 0 (dose 1), 21 (dose 2), and 42 following vaccination with A/H5N1 (A/Indonesia/05/2005) vaccine with AS03 (N=38) or MF59 (N=42) adjuvant or as unadjuvanted vaccine (N=50) were probed on two different influenza protein microarrays. The first array primarily contained monomeric HA (Array #1; Supplemental Table 2) and the second array consisted of both monomeric and trimeric HA (Array #2; Supplemental Table 3). Array #1 used for the first probing experiment (Exp 1) was composed of 284 antigens including 279 unique influenza HA antigens: 94 full-length HA (HA0; HA1+HA2) and 80 HA1 from group 1 (including H5) influenza viruses; and 56 HA0 and 38 HA1 subunits from group 2 influenza viruses. Among the group 1 antigens were 44 HA0 and 36 HA1 subunits from H5 viruses (the same subtype as the vaccine strain). In addition, there were 4 HA2 subunit antigens (2 x H5, 1 x FluB, 1 x H1, which contain most of the stalk, although misfolded), 4 NA, and 1 NP protein on Array #1. These proteins were all acquired from Sino Biologicals. Array #2 used in the second probing experiment (Exp 2) was composed of 270 unique influenza proteins including 239 HA proteins, representing 161 (57%) proteins which were included in Array #1 (Sino Biologicals), plus 66 HA0 trimers and 22 NA proteins, provided by the Dr. F. Krammer laboratory and Dr. J. Crowe laboratory. Array #2 contained 95 HA0 and 45 HA1 subunits from group 1 influenza viruses and 69 HA0 and 17 HA1 subunits from group 2 influenza viruses. Among the group 1 antigens were 50 HA0 and 33 HA1 subunits from H5 viruses. In addition, 2 HA2 antigens from H5 viruses, 1 NP, and 30 NA proteins were present on Array #2.

**Figure 1.**
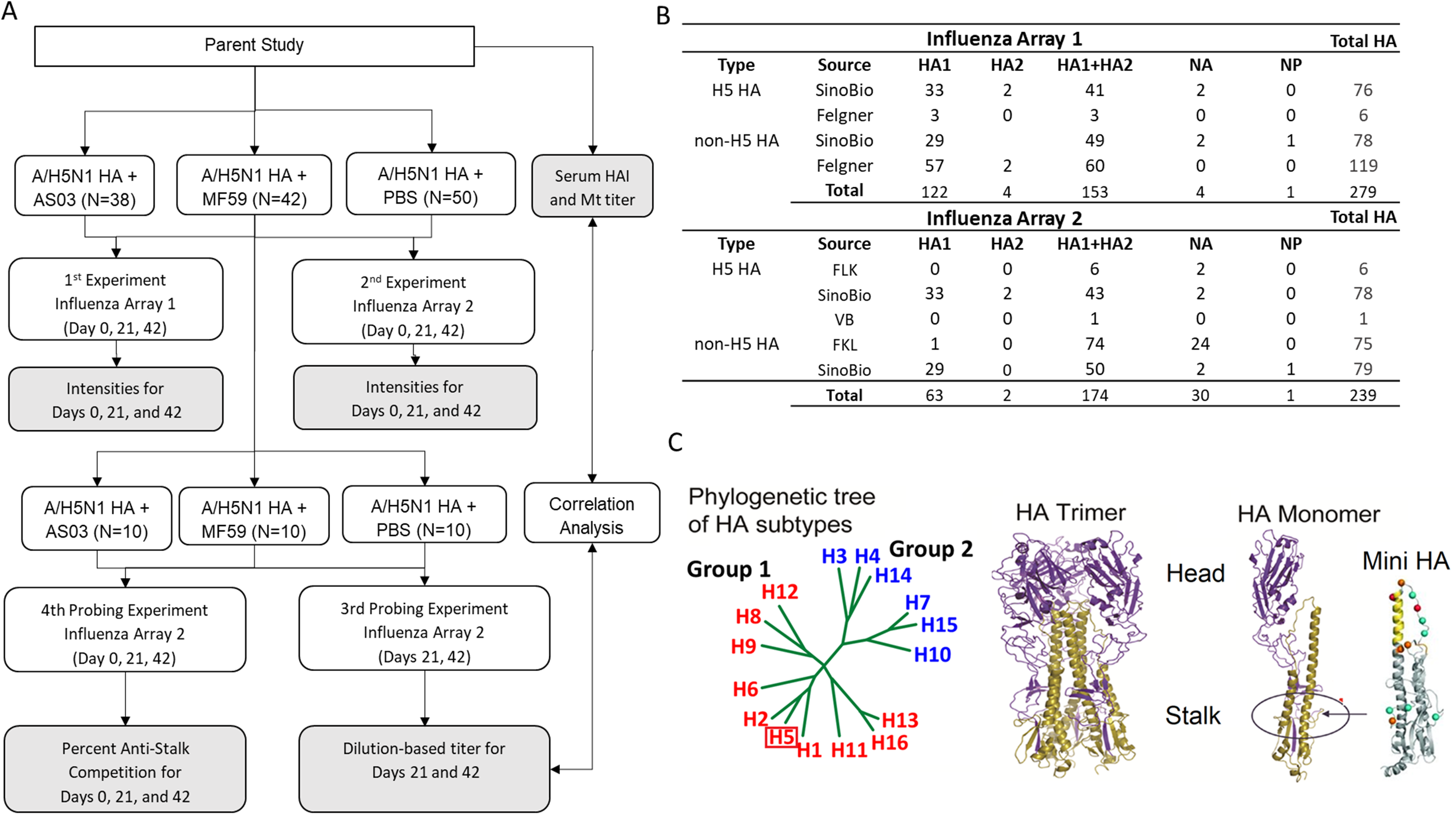
Study Overview. A: Study design B: Protein array antigen composition C: HA Subtype relationship, H5 trimer structure, MiniHA binding used for assessing percent anti-stalk competition.

### Pre-vaccination reactivity against influenzas viruses and the impact of prior seasonal influenza vaccination

A birds’ eye overview of all IgG and IgA results for both arrays and antibody isotypes (IgG and IgA) is shown in Figure 2. To bring out key signals, log_2_ fluorescent intensities for each antigen were scaled across all samples by subtracting the mean and dividing by the standard deviation (Z-score). The heatmaps show that pre-vaccination (day 0) reactivity patterns for each of the 3 vaccine groups were similar. For both Array #1 and Array #2, pre-vaccination IgG and IgA antibodies were highest against seasonal influenza virus antigens (H1, H3, or FluB) HA0 and HA1 (Supplemental Figure S1), with higher responses against HA0 than HA1. While H5 antibodies ranked in the lower third for HA1 responses, their responses ranked slightly lower than seasonal virus antigens based on HA0 responses. We inferred that these reactivities against H5 HA0 as caused by cross-reactive antibodies were originally induced by seasonal H1 exposure, as these were presumed H5-naïve subjects with no overt reason for prior exposure to H5 influenza. This prompted us to explore the impact of prior seasonal influenza vaccination on the microarray antibody responses. Exploratory inspection of covariates showed that subjects reporting seasonal influenza vaccination within the prior two years resulted in lower fold changes in antibody intensities against vaccine subtype antigens (H5 HAs) relative to pre-vaccination irrespective of experiment, vaccine group, antibody type or timepoint (Supplementary Figure S2). Sex and age did not have a large impact on fold changes.

**Figure 2.**
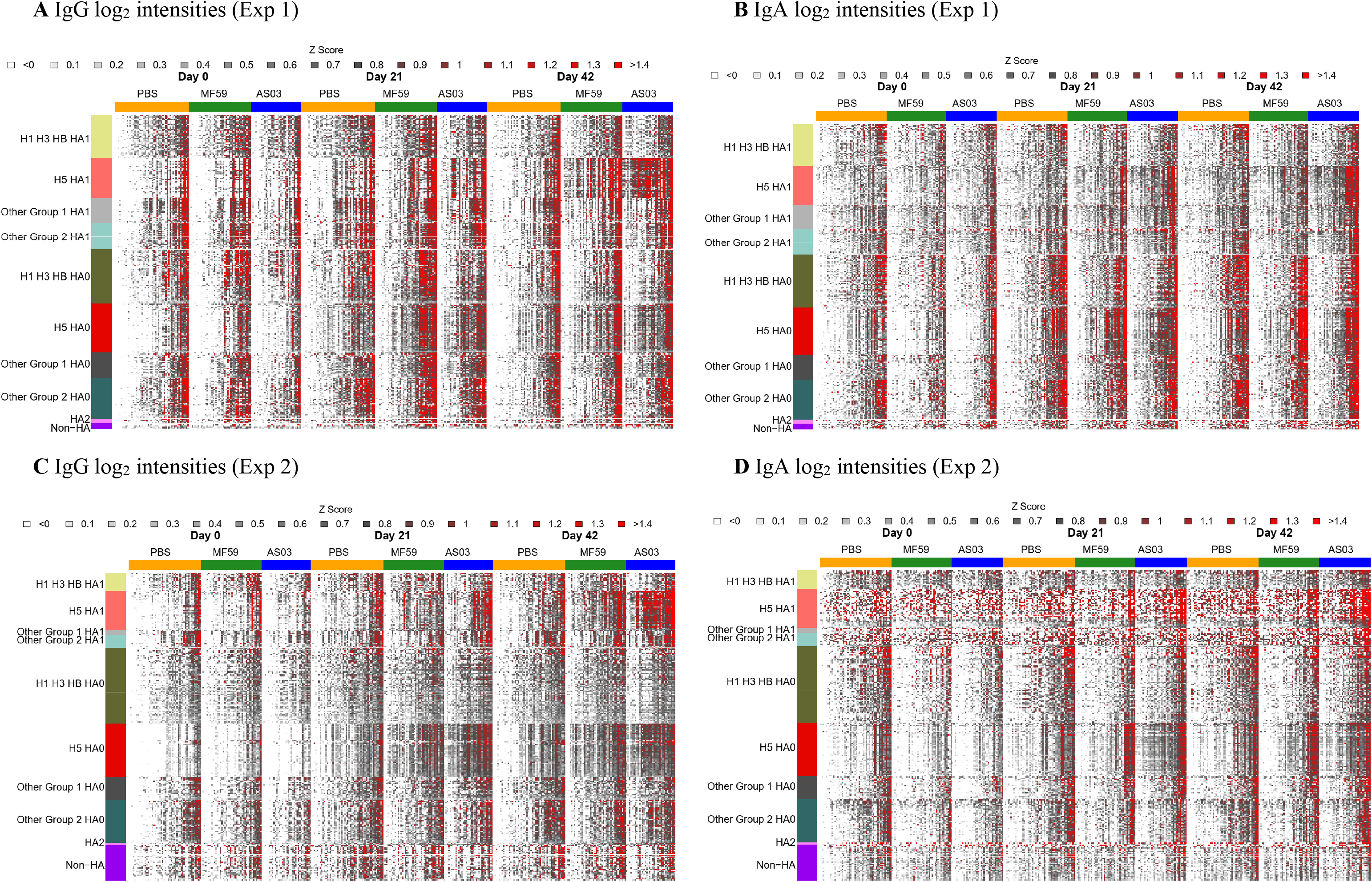
Heatmaps summarizing experiment 1 & 2 protein array IgG and IgA results by vaccine group, day, HA molecule, and HA subtype. A and B: log_2_ fluorescent intensity results by HA subtype for Experiment 1 IgG and IgA antibodies, respectively. C and D: Corresponding results for Experiment 2. Each column represents one sample from a subject, and each row represents an antigen. The antigens are organized into groups, headgroup (HA1) or full-length molecule (HA0). The HA subtypes were grouped as H1/H3/FluB, H5, Other Group 1, and Other Group 2. Fluorescent intensities were background corrected, log_2_ transformed, and standardized across rows (z-score, mean=0, variance=1). In red: intensities strongly increased compared to the mean log_2_ intensity, in dark grey: moderately increased compared to the mean in white decreased relative to the mean. Samples were ordered by increasing reactivity from left to right.

### MF59 and AS03 adjuvant effect on the homosubtypic response

To assess the strength and breadth of homosubtypic antibody responses against H5, we plotted the H5 antigens in order from the most to the least reactive (Figure 3). Again, all the pre-vaccination (day 0) results for the three vaccine groups overlapped indicating that baseline antibody levels were very similar (Figure 3 A Exp1 and B Exp 2), as assessed between the two arrays. The adjuvanted vaccines showed higher geometric mean intensity (GMI) after the first dose (day 21) and even higher GMI after the second dose of adjuvanted vaccine (day 42) compared to unadjuvanted vaccine. The relative antibody induction levels measured in both experiment 1 and 2 were AS03>MF59>unadjuvanted. To better display differences between doses for each adjuvant, we also presented the data within each vaccine group by day (Figure 3 C Exp 1 and D Exp 2). In both experiments, the unadjuvanted vaccine induced IgG against H5 HA0 on day 21 after the first dose that was not significantly boosted further on day 42 (21 days after the second dose). There was much weaker induction of IgG by the unadjuvanted vaccine on day 21 or 42 against HA1 and no substantial IgA response against the HA1 or HA0 antigens at either time point. Both the MF59 and AS03 adjuvanted vaccine induced IgG against HA1 after the first dose and increased after the second dose; however, the IgG response against HA0 did not increase after the second dose. The MF59 and AS03 adjuvants also induced IgA against HA0 that also did not increase after the second dose.

**Figure 3.**
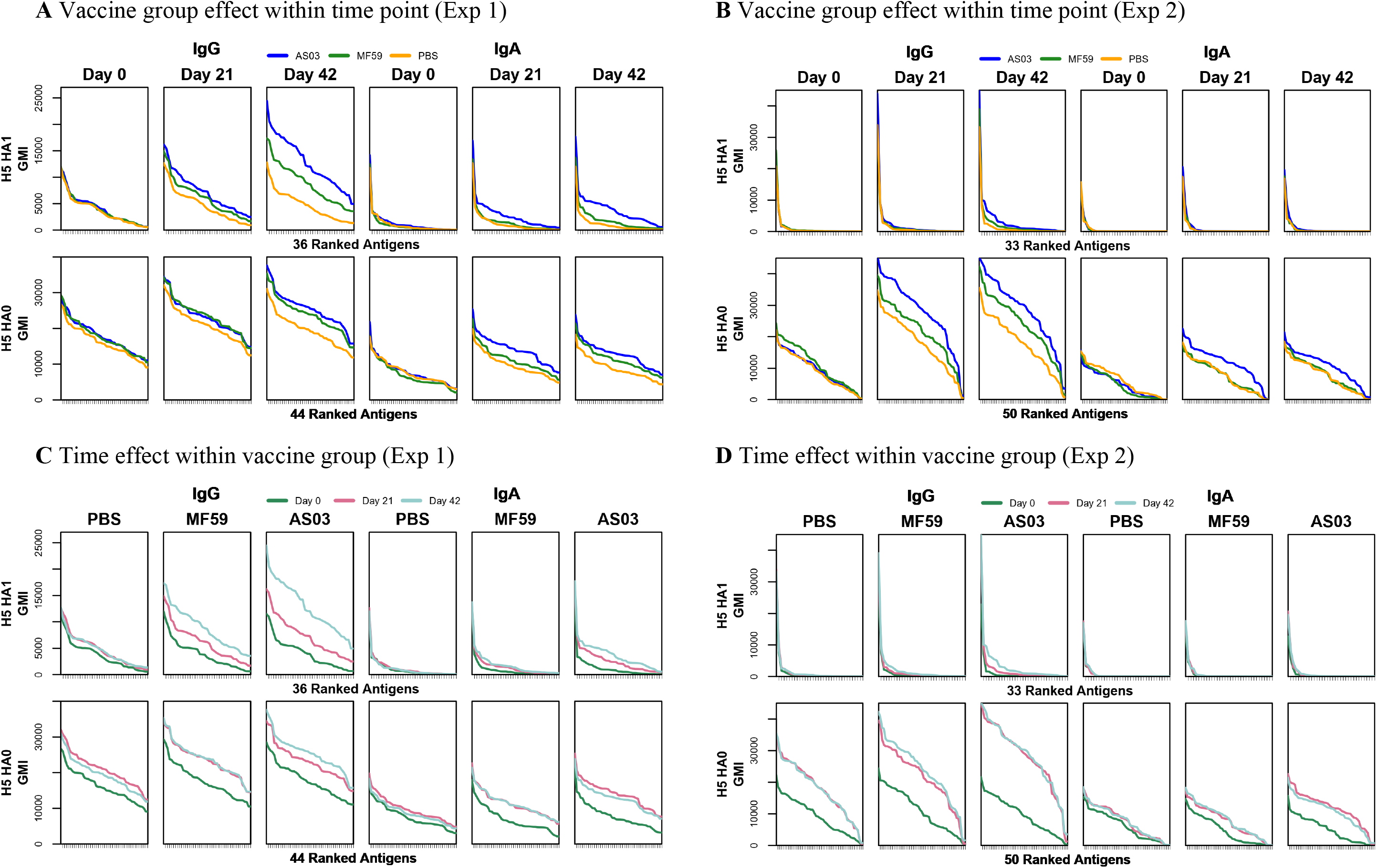
Time trend plots of ranked IgG and IgA antibody responses against H5 HA1 and HA0 antigens based on geometric mean fluorescent intensity over time. A, C: Experiment 1 results by vaccine group following each dose; B, D: Experiment 2. HA1: antibody responses against head H5 HA proteins, HA0: antibody responses against full length H5 HA proteins. GMI: Geometric mean fluorescent intensity.

### Adjuvant mediated induction of antibodies against the HA1 and HA0 molecule

Next, we analyzed adjuvanted vaccine responses after adjusting for pre-vaccination level and prior influenza vaccination. The mean fold change relative to pre-vaccination adjusted for seasonal influenza vaccination is summarized in Figure 4. The analysis compares subtype-specific antibody levels induced by the three vaccine groups. Compared to adjuvanted vaccines, the unadjuvanted vaccine induced minimal fold changes relative to pre-vaccination among all the HA subtypes at both day 21 and day 42. In Exp 1 (Array #1) the adjuvanted vaccines induced a substantial fold increase in IgG and IgA against H5 HA1 after the first dose and there was a further increase after the second dose; these adjuvanted vaccine responses were greater for AS03 than MF59 (Figure 4A). However, the response patterns against H5 HA0 were less evident. We attribute the more profound responses to HA1 compared to HA0 to cross reactivity of antibody against the H5 stalk, which could have been elicited from prior seasonal group 1 vaccination or infection.

**Figure 4.**
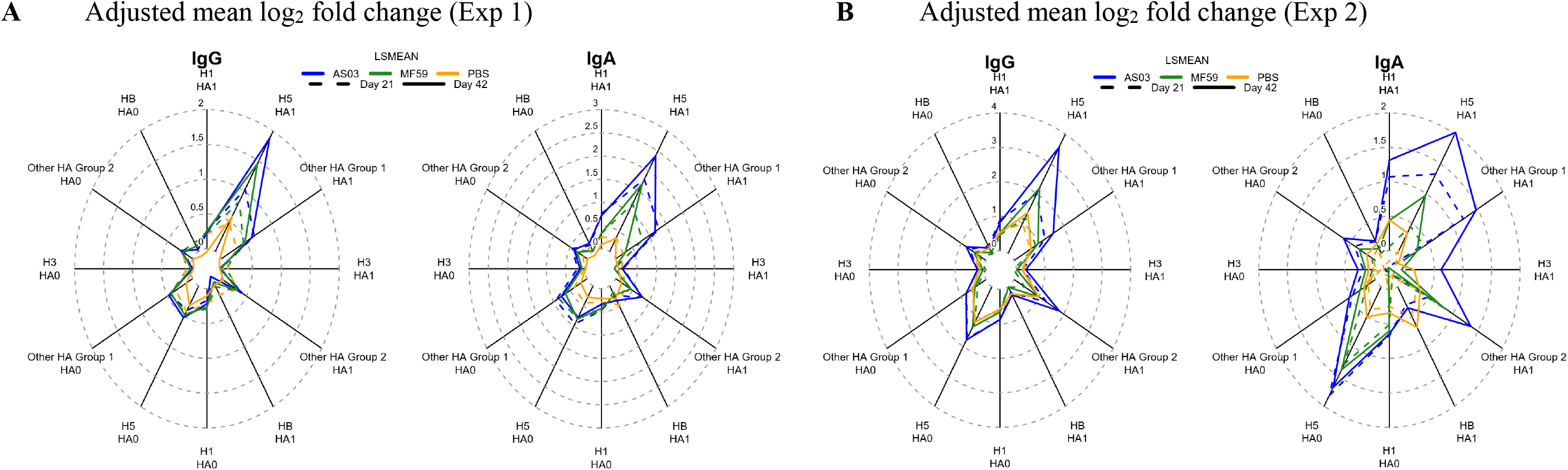
Adjuvants boosted IgG and IgA antibodies against head H5 HA proteins post-second dose. A: Experiment 1 IgG and IgA radar plots summarizing the mean log_2_ fold change relative to pre-vaccination adjusted for prior seasonal influenza vaccination. B: Corresponding Experiment 2 results. For each antigen, first the mean log_2_ fold change in antibody fluorescent intensity relative to pre-vaccination adjusted for prior seasonal influenza vaccination was determined (LSMEAN). The value for each ray in the radar plot was then calculated as the mean of the LSMEANs for the respective antigen group, vaccine group, and timepoint combination.

Array #2 for Exp 2 included 88 stabilized HA trimers not present on Array #1. Full length stabilized trimeric HA molecules detected more antibodies directed against conserved conformational epitopes presented by the trimer than monomeric HA in Array #1 (Supplementary Figure S3). In Array #2, AS03 induced an IgG and IgA response against HA1 on day 21 that increased on day 42 (Figure 4B). However, the IgA response in Array #2 against full length trimeric H5 HA for the adjuvanted groups was more pronounced than the IgA responses in Array #1 showing an increase on day 21 that did not increase on day 42. This difference between Array #1 and #2 may be attributed to the presence of additional conformational epitopes present in the stabilized trimers. This IgA heterosubtypic cross-reactive response against group 1 HA was evident from the adjuvanted vaccines that was not boosted with a second dose, which we interpret as indicating heterosubtypic cross reactivity against the conserved stalk. It is possible the heterosubtypic group 1 HA1 responses induced by the adjuvanted vaccines were to the N-terminus of HA1, which contains a conserved 115 amino acid sequence that forms part of the cross-reactive stalk (Figure 1C).

### Adjuvant effect on heterosubtypic responses

To statistically assess the impact of the adjuvants on heterosubtypic breadth of coverage, we fit ANOVA models for day 21 and 42 comparing adjuvanted vs. unadjuvanted responses adjusting for pre-vaccination and prior seasonal influenza vaccination (Table 1). For Array #1, at day 42 (21 days post-second dose), AS03-adjuvanted vaccine elicited a higher number of significant IgG responses and a broader coverage of significant responses against non-H5 HAs compared to MF59. This was demonstrated by a significant increase in IgG responses against 78 unique HA antigens in the AS03 group (of which 47% were non-H5 HA antigens) and 37 unique HA antigens (among which are a subset of the 78 HA antigens recognized in the AS03 group) in the MF59 group (22% non-H5 HA antigens) compared to the unadjuvanted group. This difference in the recognition of non-H5 HA antigens was statistically significant (Fisher’s exact test < 0.01) with the AS03 group eliciting more responses against non-H5 HA0 antigens (Table 1). For both adjuvanted groups, the most frequent responses relative to the unadjuvanted group were IgG responses against HA1 antigens (AS03: 73%, MF59: 89%), and these were predominantly among H5 HA1 antigens (AS03: 63%, MF59: 82%). When assessing IgA responses in Array #1, the AS03 group showed statistically significant responses against 28 unique antigens at day 21 and 37 antigens at day 42. Most of these AS03 responses (76%) were against HA0 at day 21, of which 43% were against non-H5 HA0; at day 42, 61% of the unique antigens were against HA0, of which 49% were against non-H5 HA0. A substantial additional number of H5 HA1 antigens were recognized by IgA at day 42 (30%) relative to day 21 (16%). In contrast to IgG, there were more IgA responses against HA0 with AS03 at day 42 (Fisher’s exact test < 0.001).

**Table 1.** Summary of statistically significant adjuvant-boosted antibody responses. For each antigen, an ANOVA model was fitted to day 21 and day 42 log_2_ fold change responses to assess the statistical significance of contrasts that compared adjuvanted vs. control group responses (AS03 - PBS and MF59 - PBS) adjusting for prior seasonal influenza vaccination using a FDR-adjusted p-value < 0.1.

|  |  |  | Experiment 1 |  |  |  |  |  |  |  | Experiment 2 |  |  |  |  |  |  |  |
| --- | --- | --- | --- | --- | --- | --- | --- | --- | --- | --- | --- | --- | --- | --- | --- | --- | --- | --- |
|  | Day | Subtype | IgG |  |  |  | IgA |  |  |  | IgG |  |  |  | IgA |  |  |  |
| Comparison | Day | Subtype | HA1 | HA0 | Total | % Total | HA1 | HA0 | Total | % Total | HA1 | HA0 | Total | % Total | HA1 | HA0 | Total | % Total |
| AS03 vs. PBS | 21 | H5HA | 0 | 0 | 0 | 0 | 6 | 16 | 22 | 59 | 0 | 0 | 0 | 0 | 4 | 32 | 36 | 0 |
|  |  | non-H5HA | 0 | 0 | 0 | 0 | 3 | 12 | 15 | 41 | 0 | 0 | 0 | 0 | 4 | 17 | 21 | 0 |
|  |  | Total | 0 | 0 | 0 | 0 | 9 | 28 | 37 |  | 0 | 0 | 0 | 0 | 8 | 49 | 57 | 0 |
|  | 42 | H5HA | 36 | 5 | 41 | 53 | 18 | 19 | 37 | 61 | 23 | 8 | 31 | 82 | 7 | 23 | 30 | 79 |
|  |  | non-H5HA | 21 | 16 | 37 | 47 | 6 | 18 | 24 | 39 | 1 | 6 | 7 | 18 | 0 | 9 | 9 | 24 |
|  |  | Total | 57 | 21 | 78 |  | 24 | 37 | 61 |  | 24 | 14 | 38 |  | 7 | 32 | 39 |  |
| MF59 vs. PBS | 21 | H5HA | 0 | 0 | 0 | 0 | 0 | 0 | 0 | 0 | 0 | 0 | 0 | 0 | 0 | 0 | 0 | 0 |
|  |  | non-H5HA | 0 | 0 | 0 | 0 | 0 | 0 | 0 | 0 | 0 | 0 | 0 | 0 | 0 | 0 | 0 | 0 |
|  |  | Total | 0 | 0 | 0 | 0 | 0 | 0 | 0 | 0 | 0 | 0 | 0 | 0 | 0 | 0 | 0 | 0 |
|  | 42 | H5HA | 27 | 2 | 29 | 78 | 8 | 19 | 27 | 64 | 0 | 0 | 0 | 0 | 0 | 0 | 0 | 0 |
|  |  | non-H5HA | 6 | 2 | 8 | 22 | 3 | 12 | 15 | 36 | 0 | 0 | 0 | 0 | 0 | 0 | 0 | 0 |
|  |  | Total | 33 | 4 | 37 |  | 11 | 31 | 42 |  | 0 | 0 | 0 |  | 0 | 0 | 0 |  |

For the MF59 group, 42 statistically significant IgA responses were identified, of which 31 (74%) were against HA0 and from which 39% were against non-H5 HA0. In contrast to IgG, the difference in H5 vs. non-H5 HA antigen recognition at day 42 between the AS03 and MF59 group was not statistically significant. In Array #2, only the AS03 adjuvant was associated with statistically significant responses that differed compared to the unadjuvanted group (Table 1). This included IgG against 38 antigens, of which 31 (82%) were H5 HA and 7 (18%) were non-H5 HA antigens. While most H5 HA IgG responses were observed against HA1 (74%), responses against non-H5 HA were primarily observed against HA0 (86%). In addition, the AS03 group elicited statistically significant IgA against 57 antigens at day 21 and 39 antigens at day 42. The majority of these responses were against HA0 antigens, of which 35% were against non-H5 HA0 at day 21 and 28% at day 42.

### Adjuvant effect on the antibody landscape of group 1 and 2 influenza virus antigens

Next, to further assess heterosubtypic breath, we generated antibody landscapes that profile the antibody response against influenza virus antigens as a function of antigenic differences on the sequence level (Figure 5). The landscapes contrast homosubtypic and heterosubtypic breadth and intensity of the IgG and IgA response against the 279 HA variants elicited by the three vaccine groups separately, against either the HA1 (Exp 1: Figure 5A and B) or the HA0 molecules (Exp 2: Figure 5C and D) and are adjusted for prior-influenza vaccination. On day 21, AS03 adjuvanted vaccine induced homosubtypic responses against H5 HA1 variants and heterosubtypic cross-reactive responses against closely related group 1 antigens that increased on day 42 (Figure 5A, Figure 5B). A similar pattern was seen with MF59, but the effect was less pronounced than for AS03. In contrast, the response with unadjuvanted vaccine, while still showing a distinct homosubtypic response against H5 HAs, was much weaker. Antibody landscapes for IgG against HA0 are shown in Figure 5C. In the adjuvanted vaccine groups, the IgG responses after a second dose (day 42) with HA1 (Figure 5A and Figure 3) were not seen against HA0 (Figure 5C). However, all three vaccine groups induced increased IgG against H5 HA0 proteins with similar intensities after one and two doses (Figure 5C). For IgA, in contrast to IgG, the higher response to a second dose was only seen in the adjuvanted groups and was absent from the unadjuvanted group (Figure 5D).

**Figure 5A.**
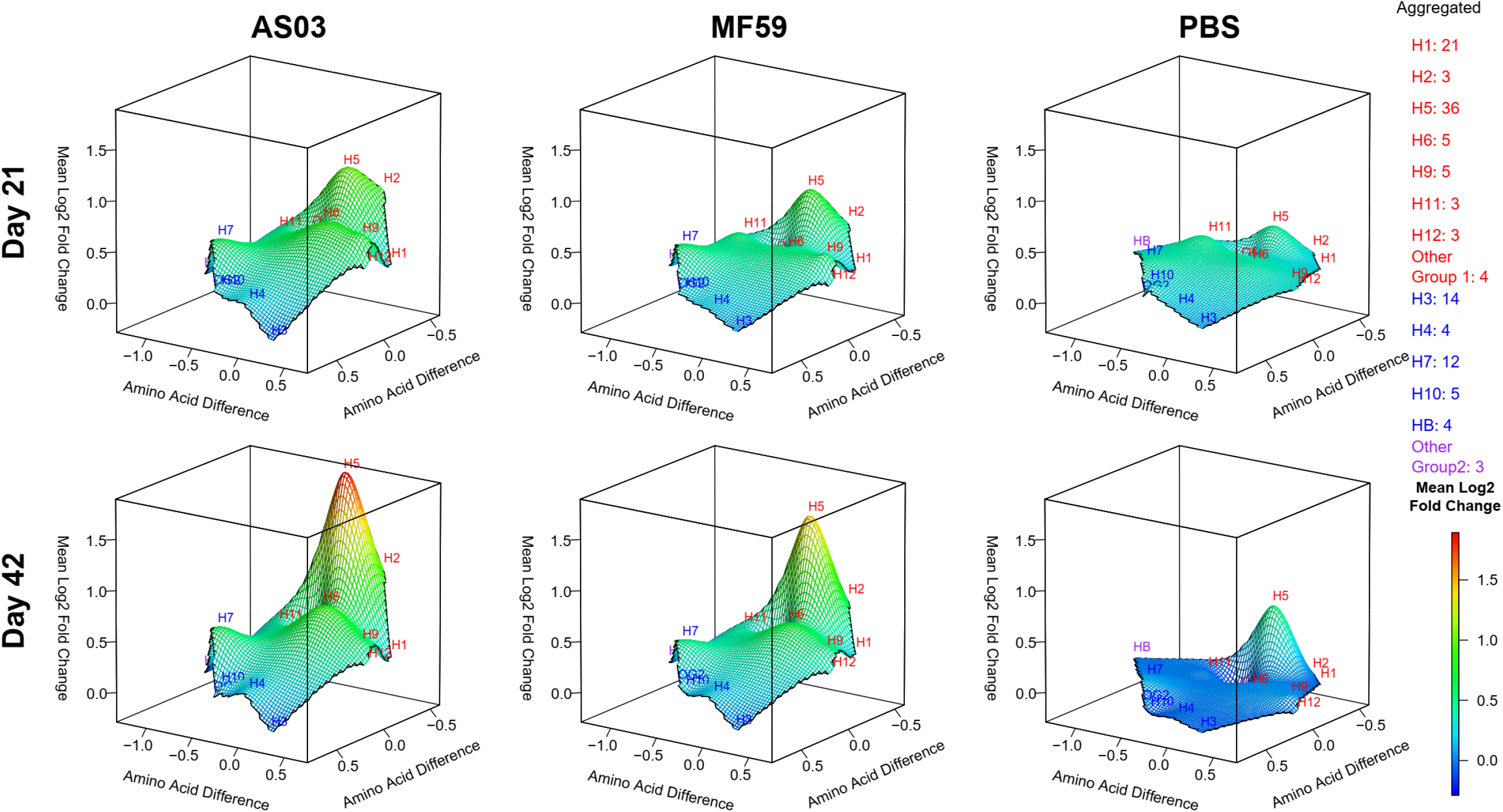
Adjuvants boosted IgG antibodies against head H5 HA proteins post-second dose. IgG antibody landscapes for HA1 subunit antigens based on mean log_2_ fold change relative to pre-vaccination adjusted for seasonal influenza vaccination (Experiment 1). In red: Group 1 HA1 antigens, in blue: Group 2 HA1 antigens, in purple: influenza B HA1. The y and x axes project antigen protein sequence divergence as measured in the number of amino acid differences between antigen protein using multidimensional scaling. For each antigen, first the mean log_2_ fold change in antibody fluorescent intensity relative to pre-vaccination adjusted for prior seasonal influenza vaccination was determined (LSMEAN). For subtypes with multiple antigens, the mean LSMEAN and MDS dimensions were used to present the results in the 3D space.

**Figure 5B.**
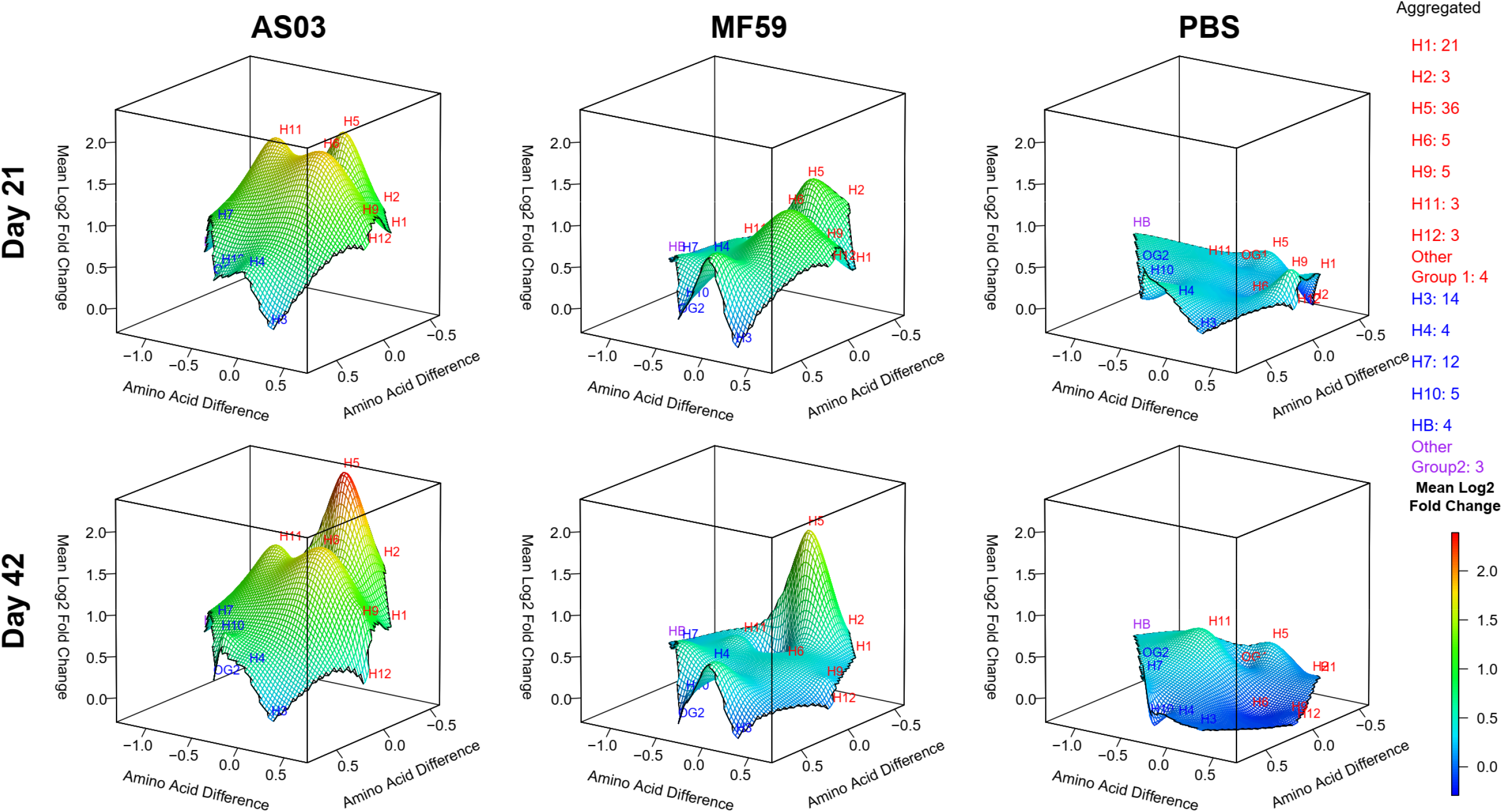
Adjuvants boosted IgA antibodies against head H5 HA proteins post-second dose. IgA antibody landscapes for HA1 subunit antigens based on mean log_2_ fold change relative to pre-vaccination adjusted for seasonal influenza vaccination (Experiment 1). In red: Group 1 HA1 antigens, in blue: Group 2 HA1 antigens, in purple: influenza B HA1. The y and x axes project antigen protein sequence divergence as measured in the number of amino acid differences between antigen protein using multidimensional scaling. For each antigen, first the mean log_2_ fold change in antibody fluorescent intensity relative to pre-vaccination adjusted for prior seasonal influenza vaccination was determined (LSMEAN). For subtypes with multiple antigens, the mean LSMEAN and MDS dimensions were used to present the results in the 3D space.

**Figure 5C.**
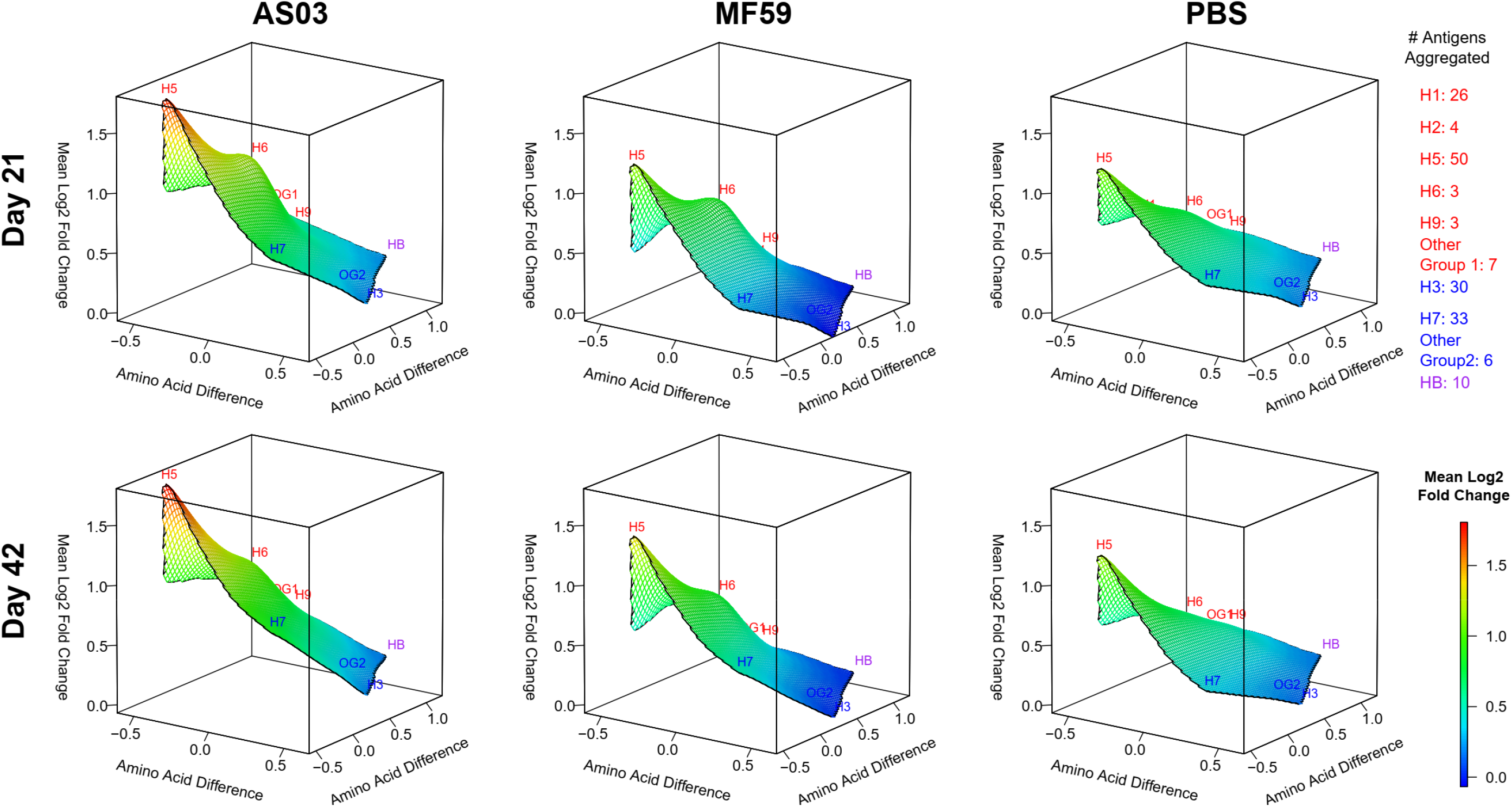
All three vaccines elicited increased IgG antibodies against full-length H5 proteins after the first dose without further boosting after the second dose. IgG antibody landscapes for HA1 subunit antigens based on mean log_2_ fold change relative to pre-vaccination adjusted for seasonal influenza vaccination (Experiment 2). In red: Group 1 HA1 antigens, in blue: Group 2 HA1 antigens, in purple: influenza B HA1. The y and x axes project antigen protein sequence divergence as measured in the number of amino acid differences between antigen protein using multidimensional scaling. For each antigen, first the mean log_2_ fold change in antibody fluorescent intensity relative to pre-vaccination adjusted for prior seasonal influenza vaccination was determined (LSMEAN). For subtypes with multiple antigens, the mean LSMEAN and MDS dimensions were used to present the results in the 3D space.

**Figure 5D.**
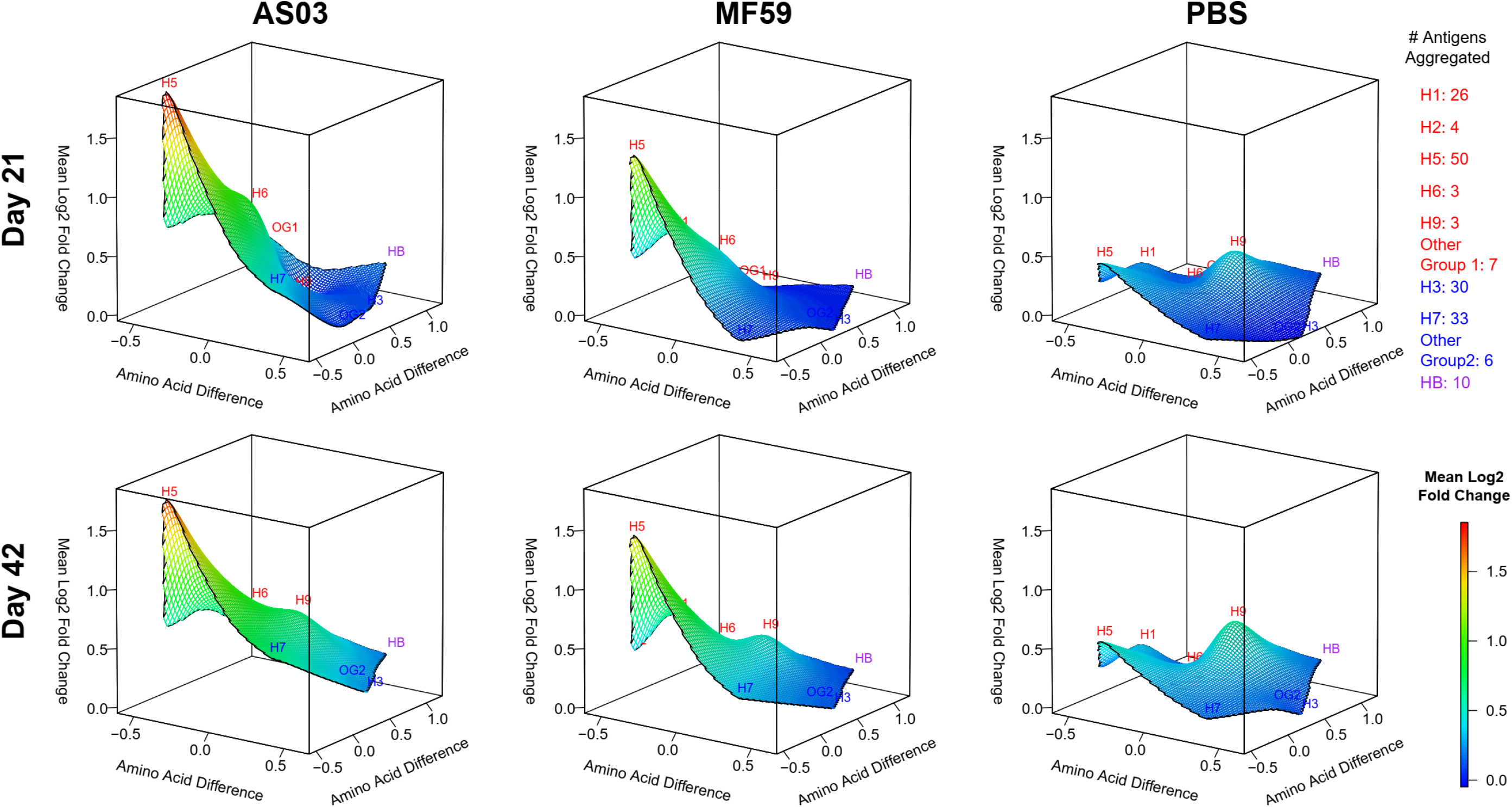
Only adjuvanted vaccines elicited increased IgA antibodies against full-length H5 proteins after the first dose. IgG antibody landscapes for HA1 subunit antigens based on mean log_2_ fold change relative to pre-vaccination adjusted for seasonal influenza vaccination (Experiment 2). In red: Group 1 HA1 antigens, in blue: Group 2 HA1 antigens, in purple: influenza B HA1. The y and x axes project antigen protein sequence divergence as measured in the number of amino acid differences between antigen protein using multidimensional scaling. For each antigen, first the mean log_2_ fold change in antibody fluorescent intensity relative to pre-vaccination adjusted for prior seasonal influenza vaccination was determined (LSMEAN). For subtypes with multiple antigens, the mean LSMEAN and MDS dimensions were used to present the results in the 3D space.

According to the radar plots (Figure 4), which plotted the mean log2 fold change relative to pre-vaccination, there were detectable antibody responses against group 2 HA proteins with adjuvanted vaccination. These responses were more prominent among HA1 than for HA0 and were more evident among IgA than IgG antibodies. We assume the study participants were likely to have previously been primed against H3 with prior seasonal influenza vaccination or infection. The antibodies elicited by adjuvanted H5 vaccine appeared to also react with non-H3 group 2 HA proteins. However, according to the landscape plots (Figure 5), these group 2 antibody responses are minor in comparison to the magnitude of the antibody responses against homo-subtypic and hetero-subtypic HAs.

### Antibody directed against HA stalk

In order to estimate the relative levels of anti-stalk responses elicited by the 3 vaccine groups, we used a stabilized trimeric Mini H1 HA recombinant stalk molecule^20, 21, 22^ (#4900), produced by Dr. L. Coughlan laboratory), which lacks the entire HA1 head including the HA1 segment that interacts with the stalk. As a negative control we used a pre-fusion stabilized trimeric RSV F protein (DS-Cav1 5K6I, also produced by Dr. L. Coughlan laboratory)^23^. We performed anti-stalk experiments for a subset of sera from 30 subjects (10 per vaccine group). Trends of anti-stalk blocking effects by Mini H1 HA concentration (0, 10, and 50 μg/mL) are provided in Supplemental Figures 4 and 5.

The pre-incubation of mini-HA with serum specimens reduced up to 65% of the IgG and 67% of the IgA antibody binding responses detected on Array #2 relative to the negative control when using the intensity area under the curve for both concentrations (Figure 6). We attribute this reduction in detected binding responses to competitive binding of the mini-HA to anti-stalk specific antibody in the sample. Baseline (pre-vaccination) specimens demonstrated up to 25% diminishment of the H1 HA0 IgG and IgA signal with mini-HA pre-treatment. After vaccination (days 21 and 42), the percentage of H1 HA0 IgG and IgA blocked by the mini-HA increased, consistent with an increase in vaccine induced heterosubtypic cross-reactive antibody responses against H1. From baseline specimen, 50% of the H5 IgG signal was blocked by mini-HA, indicating that a large portion of the pre-existing IgG response in the H5 signal was directed against the stalk region of HA. Since the study participants were enrolled using criteria to minimize the risk of prior exposure to H5 viruses or H5 vaccination, we infer that the baseline H5 responses are likely those elicited by cross-reactive antibodies from prior seasonal H1 exposure and/or vaccinations.

**Figure 6.**
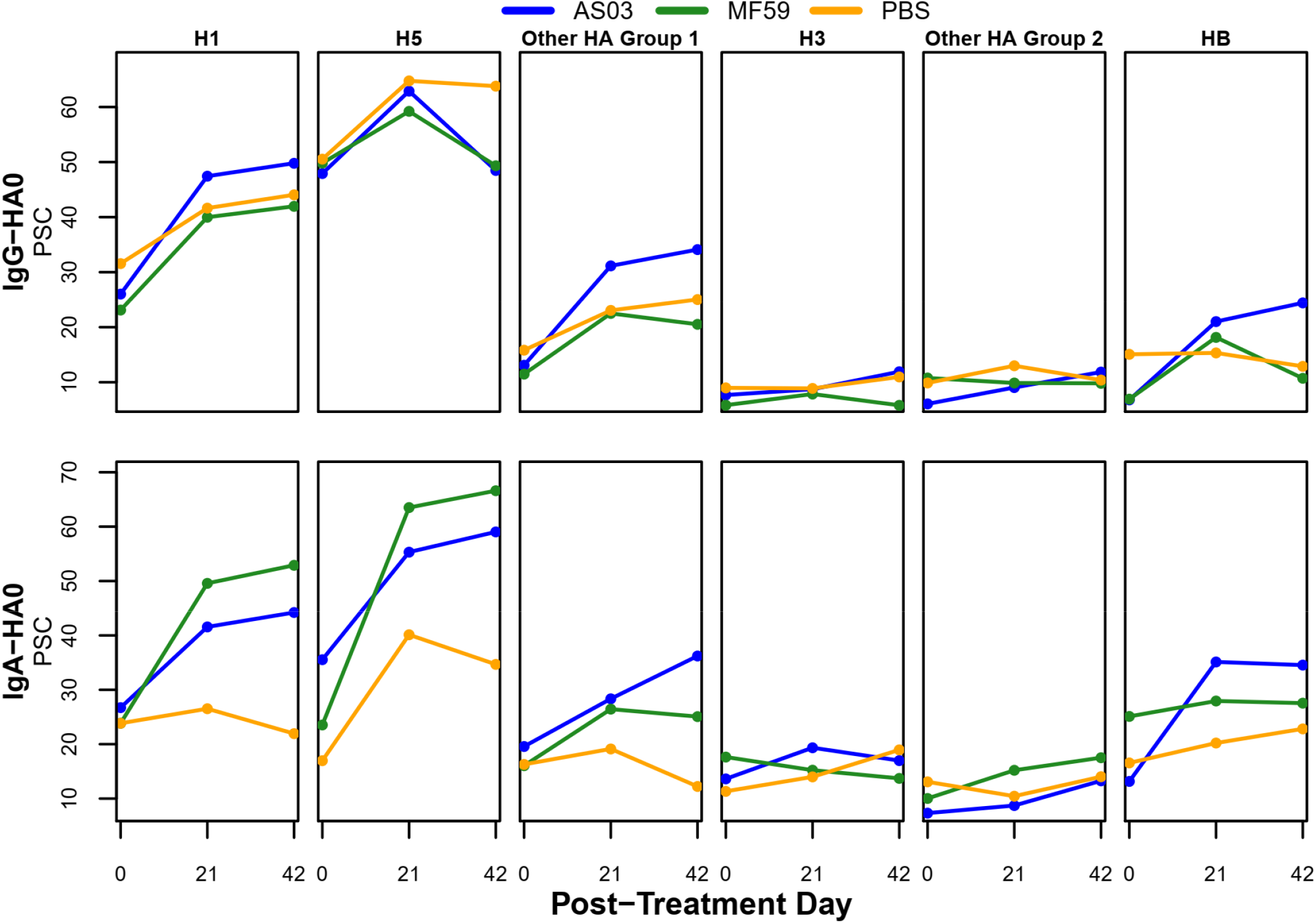
Median percent anti-stalk blocking results for IgG and IgA antibodies against full length HA antigens over time. Percent stalk competition (PSC) was calculated as: 100 - (AUC with competition (miniHA added) / AUC without competition (RSV-F negative control added) x 100) where the AUC represented the area under the curve for fluorescence intensity for two different concentrations of miniHA (10 µg and 50 µg/mL). For each vaccine group (n=10 each), the median AUC was determined. Median values were then aggregated across antigens for the respective antigen group using the median.

At day 21 (post-first dose), all three vaccine groups demonstrated the highest degree of stalk directed responses. These purported stalk responses were cross-reactive against H1, H5, and other group 1 HA antigens; the adjuvanted groups elicited much more pronounced IgA anti-stalk responses compared to the unadjuvanted group. Except for the IgG responses against H5, day 42 (post-second dose) demonstrated a further increase in anti-stalk responses, but this was weaker than the increase after the first dose. On the other hand, the day 42 IgG purported stalk antibody response was lower than that at day 21. We hypothesize this was the result of the majority of the antibody response being directed against the immunodominant head group of H5 subsequent to the second dose of vaccine. Other group 1 HA molecules responded in a similar pattern to those of the H1 and H5 responses, which we attribute to anti-stalk directed responses post vaccination. Interestingly, AS03 adjuvanted H5 vaccination was also associated with an increase in anti-stalk antibody.

### Correlation of microneutralization and hemagglutination inhibition titers with microarray dilution-based titers

To get a more robust assessment of antibody responses, we determined titers for a subset of sera from 30 subjects (10 per vaccine group) and two timepoints (day 21 and 42). For each serum specimen, we performed eight 2-fold serial dilutions, beginning at 1/50-fold dilution, on Array #2, to generate 239 titration curves for each of the 60 samples tested (1/50, 1/100, 1/200, 1/400, 1/800, 1/1600, 1/3200, 1/6400) (Figure 7A). The titration curves generated in this way follow expected saturation curves [Ab] + [Ag] *⇌* [AbAg] where K_D_ = [Ab] [Ag] / [AbAg]. Since the antigen specific concentration of antibody cannot be directly measured in polyclonal sera we substituted antibody dilution for [Ab] and used K_D_, the dilution at which the signal reached ½ of its maximal value (Bmax), as an estimate for the titer. Titers estimated from these serial dilution experiments were assessed for correlation against the known hemagglutination inhibition (HAI) and microneutralization (MN) titers in the samples. We first compared the calculated titers from the four HA0 antigens on the array that matched the vaccine HA (H5 A/Indonesia/05/2005). The IgG dilution-based titers for H5 A/Indonesia/05/2005 correlated strongly with microneutralization titers at day 21 (r_s_: 0.51 to 0.56) and 42 (r_s_: 0.57 to 0.80) and with HAI titers at day 42 (r_s_: 0.49 to 0.74). Day 42 associations for a trimeric HA0 and the cleaved version of a monomer are presented in Figure 7B. In all these cases, associations were statistically significant (p < 0.05). Whereas, the IgA dilution-based titers correlations were less strong and were more dependent on the confirmation of the vaccine antigen analyzed (HAI or MN of r_s_: 0.31 to 0.51 for day 21 and r_s_: 0.43 to 0.63) with only the trimeric HA0 reaching statistical significance at day 42 (r=0.63, p < 0.05). All day 42 associations are shown in Supplementary Figure S5.

**Figure 7.**
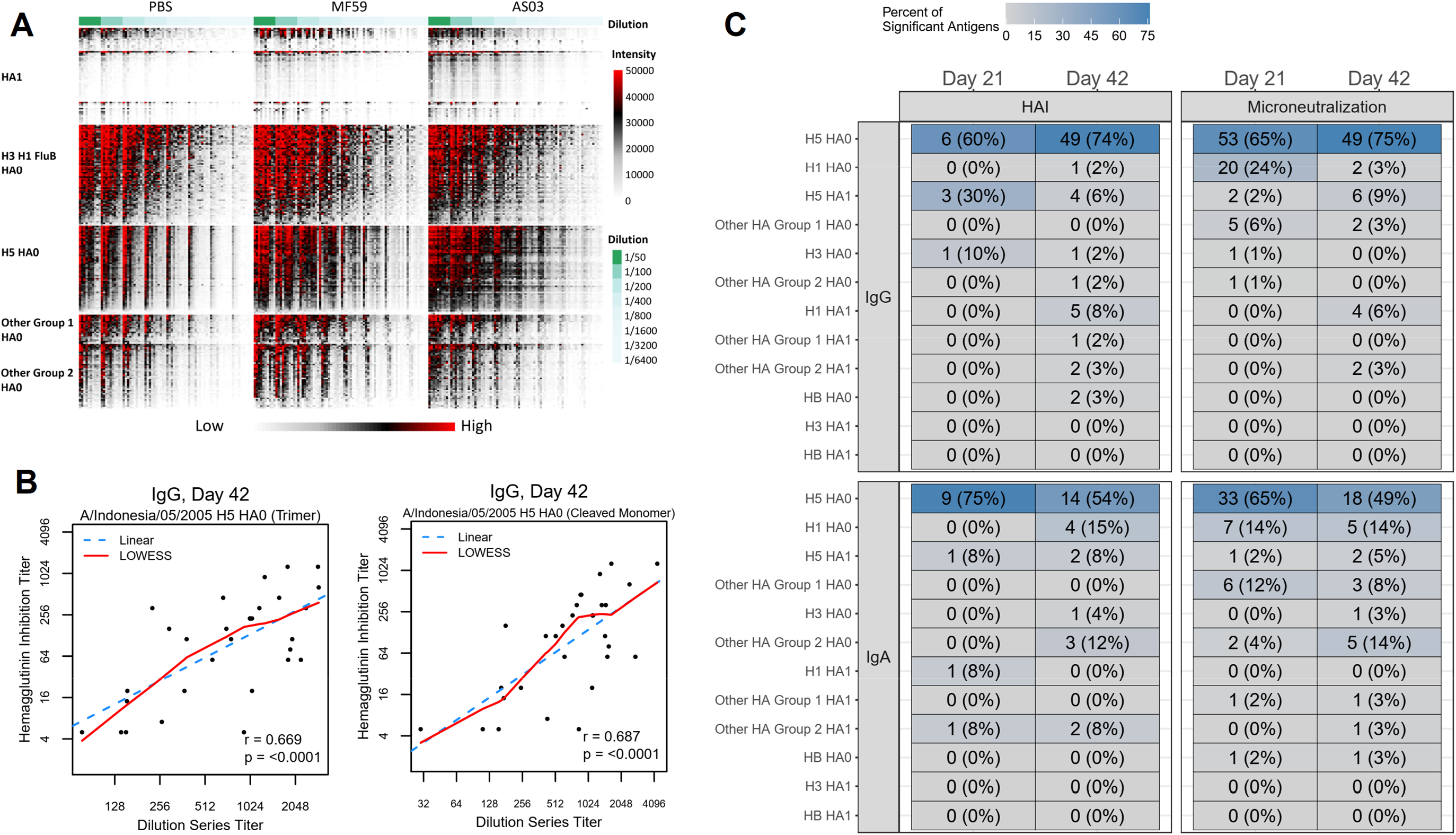
Array-based titers determined in an 8 dilution-series experiment for vaccine antigens and antigens belonging to the same subtype as the vaccine antigen were significantly correlated with HAI and microneutralization titers. A: Array results for 8 dilutions for 60 sera B: Scatter plots of day 42 log_2_ dilution series-based titer and log_2_ hemagglutinin inhibition and log_2_ microneutralization titer for influenza A/Indonesia/05/2005 antigens across vaccine groups (n=30) for HA0 trimer (left) and cleaved monomer (right), C: The number and percentage of antigens with statistically significant Pearson correlations (p < 0.05) by assay, molecule, and subtype.

Next, we assessed the correlation patterns across all antigens (Figure 7C). Assessment of statistically significant correlations (p < 0.05) across HA subtypes showed that these antigens were predominantly H5 full length antigens which were of the same subtype as the vaccine (Figure 7C). For IgG, the percentage of correlated responses among all H5 full length antigens on the array increased from 60% to 74% for HAI following the second dose indicating an increase of 14% in correlated responses. For microneutralization, an increase from 65 to 75% was observed. In contrast, IgA dilution-based titers showed the broadest correlation against H5 full length antigens at day 21 which decreased from 75% to 54 % of all H5 full length antigens on the array by HAI and 65% to 49% by MN at day 42.

## DISCUSSION

Conventional, unadjuvanted influenza virus vaccines engender antibody-mediated protection that is HA subtype-and strain-specific and relatively short-lived. Better coverage across drifted variants, or clades within a subtype, or potentially broad breadth of cross-reactivity against distinct HA subtypes could be improved with adjuvants. A better understanding of how specific adjuvants promote the elicitation of more broadly cross-reactive antibodies is needed. We constructed two sets of influenza-specific high-density protein microarrays which comprised purified HA proteins derived from 17 influenza A virus subtypes (H1 to H16 and H18) and influenza B virus strains. The arrays contained HA0 and HA1 truncated at the protease cleavage site. The arrays were used to interrogate 390 serum specimens from 130 clinical trial volunteers who received two doses of inactivated influenza A/Indonesia/05/2005 (H5N1 clade 2.2.3) virus vaccine containing 15 µg of HA with or without AS03^11^ or MF59^24^ adjuvant, administered 21 days apart.

Despite repeated exposures to seasonal influenza viruses and no known exposure to avian H5N1 viruses, our study participants’ serum exhibited antibodies with broad reactivity against all the subtypes of full-length HA represented on both arrays at baseline prior to immunization. Our data demonstrated that a large proportion of these baseline antibodies are directed against the stalk, presumably elicited from prior infection or vaccination. Prior exposure to seasonal viruses may also elicit antibody against the N-terminus of HA1, containing a conserved 115 amino acid sequence. These N-terminus HA1 antibody responses would not be diminished with the mini-HA blocking experiments. Finally, these broadly reactive antibodies were not evaluated for binding affinity to each respective HA nor can we provide a functional assessment of these antibodies in their ability to neutralize virus. So, we cannot imply the degree to which these antibodies may provide any level of protection against influenza viruses. Nonetheless, even the traditionally-used HAI antibody is not a good correlate for predicting protection against influenza infection for all ages.^25^ These baseline antibodies were of both IgG and IgA and we conjecture that parenteral vaccination elicits an IgG response, so the pre-existing IgA is more likely a reflection of prior natural infection which through a mucosal surface would elicit both IgA and IgG. These baseline antibodies are consistent with our previous findings with a similar influenza microarray study.^26^

Our results confirmed that the A/Indonesia/05/2005 H5N1 vaccine when given without an adjuvant induced limited antibody responses. This is likely a combination of the poor immunogenicity of avian HAs as well as the lack of innate immune signaling induced by adjuvants that enhances antigen processing and presentation.^27, 28^ The poor immunogenicity of unadjuvanted avian influenza vaccines is well documented.^27^ One theory is that the head domains of avian (i.e. H5) HAs are so antigenically distinct from seasonal H1, that the head lacks available T cell epitopes which could provide CD4 T cell help/Tfh to B cells.^29, 30^ The germinal center response is capable of producing IgA response when the correct innate signals from the adjuvanted vaccine are present.

Two doses of the monovalent H5N1 vaccine elicited IgG and IgA which recognized a broad range of H5 subtype and group 1 specific HA antibodies.^31–34^ Adjuvanted vaccine resulted in higher antibody levels and greater breadth. The “potency” of the responses elicited were in the descending order, AS03 > MF59 > unadjuvanted. Despite the lack of exposure to H5 virus, responses were present against full length H5 at baseline, suggesting cross reactivity against the conserved stalk region of HA, the H5 HA1 response was low at baseline, slightly increased after the first dose of vaccine (prime), and significantly increased after the second dose (boost) in the context of an adjuvant, most markedly with AS03. In contrast, IgG and IgA responses against H5 HA0 increased after the first dose but were not significantly further increased with the second dose and the H5 HA0 response to the first dose was much more pronounced for IgA than for IgG. To account for this differential boosting response, we speculate that the H5N1 HA1 is a novel variant antigen that has not been previously exposed to the immune system. The priming dose is necessary to produce memory response which is evident as the anamnestic response measured after the boost. Conversely, limited responses for H5N1 HA0 which is conserved across subtypes, maybe due to their anti-stalk prime and memory from prior seasonal exposures. The anti-stalk antibody response is anamnestic after the first dose drawing on the memory pool and consequently no further increase is seen after the boost. At baseline, about 50% of the responses against full length H5 and H1 could be blocked by competitive inhibition with an H1 stalk trimer, providing evidence that a substantial percentage of the antibody against full length H5 was direct against the highly conserved stalk region.^31–34^

Sixty specimens from this study were serially diluted 8 times to produce titration curves. Midpoint titers estimated from these serial dilution experiments were assessed for correlation with the HAI and MN titers. Statistically significantly correlated antigens were present in predominantly H5 full length antigens of the same subtype as the vaccine. The fact that microarray-based dilution titers for vaccine antigens were statistically significantly associated with HAI and MN titer at day 42 lends additional confidence to these results and hints at their potential use as surrogates for HAI and MN titers.

Subtype and drift variant virus specificity experienced with the H5N1 vaccine and with seasonal influenza vaccines may be related to the low level of heterosubtypic antibodies induced by unadjuvanted vaccines and with natural infection. The use of potent immune-stimulating adjuvants demonstrated IgG and IgA levels which were higher and more broadly reactive across drifted variants and subtypes, which may correlate to improved protective efficacy that is less prone to virus variants. Even when considering vaccine mediated subtype specific responses and vaccine efficacy, unadjuvanted seasonal influenza vaccination only demonstrates 50% effectiveness.^35, 36^ The MF59 adjuvated seasonal influenza vaccine (FluAd^®^) demonstrated some improved effectiveness compared to non-adjuvanted vaccines.^37, 38^ Our data provided further evidence for both the AS03 and MF59 adjuvant as an effective means of eliciting a higher and broader antibody response compared to the unadjuvanted vaccine. While each adjuvant has previously been shown to enhance antibody responses post-vaccination^11, 24, 39^, this study, to our knowledge, is the first to comparatively assess their breadth relative to the unadjuvanted vaccine. Even though the two parallel parent clinical studies from which our specimens were derived were not originally designed to directly compare the performance of these 2 different adjuvants, the clinical protocols are sufficiently similar such that the antibody responses we describe can be generalized and compared. Although both adjuvants appeared to broaden the antibody responses predominantly to phylogenetically related HA proteins, those closely related to the vaccine strain as was evident by the antibody landscape results, AS03 elicited statistically significantly broader heterosubtypic antibody responses, compared to MF59 recognizing more non-H5 HA antigens post-second dose.

Furthermore, anti-stalk competitive inhibition and blocking experiments indicated that some of these broad antibody responses were directed against the conserved stem region of group 1 full-length HAs. In particular, the AS03-adjuvanted group showed increased percentage in anti-stalk competition for IgG for several group 1 HA full length antigens after first and second dose relative to pre-vaccination.

Results of this study inform efforts to development of a universal influenza vaccine. While there are a number of universal vaccine candidates which are being developed using non-HA based strategies (e.g., M2, nucleoprotein, neuraminidase), a large proportion of strategies are focused on conserved regions of HA.^40–43^ Our data indicates that potent immune stimulating adjuvants can improve the antibody responses elicited by standard subvirion vaccines through eliciting more homo-and hetero-subtypic antibody. We observed that stalk directed IgG and IgA antibody was elicited with the first dose. But with the second dose of H5 vaccine, either adjuvanted or non-adjuvanted, IgG responses were directed against the head group. Whereas, IgA antibody responses against the stalk were moderately boosted with the second dose of an adjuvanted vaccine; the second dose of a non-adjuvanted vaccine elicited only head group responses. If the elicitation of stalk directed antibody were the key to protection through a universal influenza vaccine construct, then it may be essential to accompany the first dose with a potent adjuvant. Also, should a second dose of universal influenza vaccine be necessary, the same immunodominant head should not be simultaneously presented as in the first dose of the vaccine. These results appear to agree with headless stabilized stem-based vaccines^42^ or heterologous chimeric HA-based vaccine^20^ strategies. In summary, there was the demonstration of increased breadth and induction of stalk as well as head antibodies associated with AS03 relative to MF59 adjuvanted vaccination. Nonetheless, our study cannot substantiate whether there may be an increase in the clinical efficacy of vaccination with AS03 for a H5N1 pandemic vaccine over MF59.

## METHODS

### Study Specimen

The serum samples originated from future-use consenting participants of parallel avian A(H5N1)/Indonesia/05/2005 influenza vaccine mix and match studies involving MF59^24^ and AS03^11, 39^ adjuvant, also known as DMID Protocols 10-0016 and 10-0017. Only the participants that received the 15 µg dose of H5 vaccine were selected for the microarray analysis; three timepoints from each participant were selected and represent baseline (day 0 or pre-vaccination), day 21 (post-first dose of vaccine), and day 42 (21 days post-second dose of vaccine).

### Clinical trials registration

NCT01317758 and NCT01317745.

### Experiment

Each serum sample was diluted in protein array blocking buffer (Maine Manufacturing, Sanford, ME) supplemented with *E. coli* lysate (GenScript, Piscataway, NJ) to a final concentration of 10 mg/mL, and pre-incubated at room temperature (RT) for 30 minutes. Concurrently, arrays were rehydrated in blocking buffer (without lysate) for 30 minutes. Blocking buffer was removed, and arrays were probed with pre-incubated serum samples using sealed chambers to ensure no cross-contamination of sample between pads. Arrays were incubated over-night at 4 °C with gentle agitation. Arrays were then washed at RT five times with TBS-0.05% Tween 20 (T-TBS), followed by incubation with QDot®-conjugated goat anti-human IgG/ IgA diluted 1:200 in blocking buffer for 2 hours at RT. After incubation in secondary antibodies, arrays were then washed three times with TTBS and once with water. Chips were air dried by centrifugation at 1,000 g for 5 minutes and scanned on a ArrayCamTM 400-S Microarray Imaging System from Grace Bio-Labs (Bend, OR) for QDot®. Spot and background intensities were measured using an annotated grid (gal) file. For ArrayCamTM a number of different settings were used to attempt to obtain readings. Microarray spot intensities were quantified using software ArrayCamTM (Grace Bio-Labs) utilizing automatic local background subtraction for each spot. The generated signal intensity values were considered raw values.

### Data normalization

Prior to the analysis, systematic intensity differences in background reactivity as measured by phosphate buffered saline with tween 20 (PBST) were corrected. This was achieved by subtracting the median raw intensity signal for the 24 PBST intensity measurements for a certain array from the array’s non-PBST antigen intensities. Following background correction, for intensities < 1, an intensity value of 1 was imputed.

### Identification of significant adjuvant-boosted antibody responses

For each antigen, an ANOVA model was fit separately to day 21 and day 42 log_2_ fold change (LFC) responses from day 0 as part of the first and second probing experiments. The model was specified to describe the LFC in antibody signal for a certain antigen as a function of vaccine group (fixed effect with three levels: A/H5N1 HA + AS03, A/H5N1 HA + MF59, and A/H5N1 HA + PBS) and receipt of seasonal influenza vaccine during the past two years (fixed effect with two levels: yes, no). The second fixed effect was added to adjust antibody responses for seasonal influenza vaccine effects. Contrasts as implemented in the *lsmeans* R package (Version 2.27.62) were used to assess statistical significance of the mean LFC difference between the adjuvanted vaccine groups (A/H5N1 HA + AS03 and A/H5N1 HA + MF59) and the unadjuvanted control group (A/H5N1 HA + PBS) adjusted for prior receipt of vaccine (H_0_: μ_adjuvanted vaccine group_ - μ_unadjuvanted control group_ = 0, H_1_: μ_adjuvanted vaccine group_ - μ_unadjuvanted control group_ ≠ 0, on the log_2_ scale). To control for testing multiple antigens, a false-discovery rate (FDR) based on the Benjamini-Hochberg procedure as implemented in the R p.adjust function was applied separately for each post-vaccination day (day 21 and 42) and vaccine group comparison. Antibody responses with an FDR-adjusted p-value < 0.1 were deemed statistically significant.

### Determination of percent anti-stalk competition

We performed anti-stalk experiments for a subset of sera from 30 subjects (10 per vaccine group) that were probed on Array #2. Percent stalk competition (PSC) was calculated as: 100 - (AUC with competition (miniHA added) / AUC without competition (RSV-F added) x 100) where the AUC represented the area under the curve for two different concentrations of miniHA and RSV-F (10 and 50 µg/mL). The AUC was calculated using the trapezoid rule implemented in R. To prevent negative values, for PSCs that were less than 1%, a value of 1% was imputed.

### Determination of antibody titers from serial dilution experiments

Dilution experiments consisted of eight serial half-log dilutions (0.04, 0.02, 0.01, 0.005, 0.0025, 0.00125, 0.000625, 0.0003125) for day 21 and 42 specimens for a subset of 30 subjects (10 per vaccine group) that were probed on Array #2. This information was used to estimate the half maximal effective dilution, referred to as a titer. For each sample, the titer was determined using the following binding model 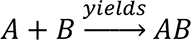 *w*here A is the antigen and B is the antibody. The functional relationship between intensity (y) and dilution (x) was described using the Michaelis-Menten equation: 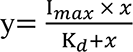. The two model parameters (I*_max_* and K*_d_*) were then estimated by fitting non-linear models as implemented in the *nls* R package. The resulting titer per sample was determined as 1/K_d_ which represented the titer estimate at which half the level of I_max_ was reached.

## Data Availability

The data will be made available following publication in a peer-reviewed journal.

## ACKNOWLEDGMENTS

We kindly thank Dr. Chris Roberts in the Respiratory Diseases Branch, Division of Microbiology/Immunology, NIAID, NIH, for facilitating access to the clinical samples and reagents and for contributions to the overall study design. We also thank Melinda Tibbals for serving as RDB project manager for this study. This study was funded through the Vaccine and Treatment Evaluation Units (VTEU) awarded to Maryland University (HHSN272201300022I and HHSN272200800057C) as well as the CRID contract awarded to the Emmes company (HHSN272200800013C and 75N93021C00012). The content is solely the responsibility of the authors and does not necessarily reflect the official views of the National Institutes of Health or the Biomedical Advanced Research and Development Authority

## ETHICS STATEMENT

Deidentified specimens, which were archived with future-use consent, were used in this study and were determined by the University of Maryland, Baltimore Institutional Review Board to consist of not human subject research.

## AUTHOR CONTRIBUTIONS

JBG conceptualized the analysis; JBG, TLJ, and SRC analyzed the data; AaJ, RA, RN, AlJ, DHD, PLF performed the experiments and consulted on the analysis and results; LC provided mini-HA and consulted on the analysis and results; JBG and WHC and PLF wrote the manuscript, WHC and PLF designed the study; CEG curated the clinical data and reviewed results, PM, DS, SS, and FK provided HA trimers and consulted on the experiment and analysis; all authors reviewed the manuscript.

## SUPPLEMENTAL FIGURES

**Supplemental Figure 1A.**
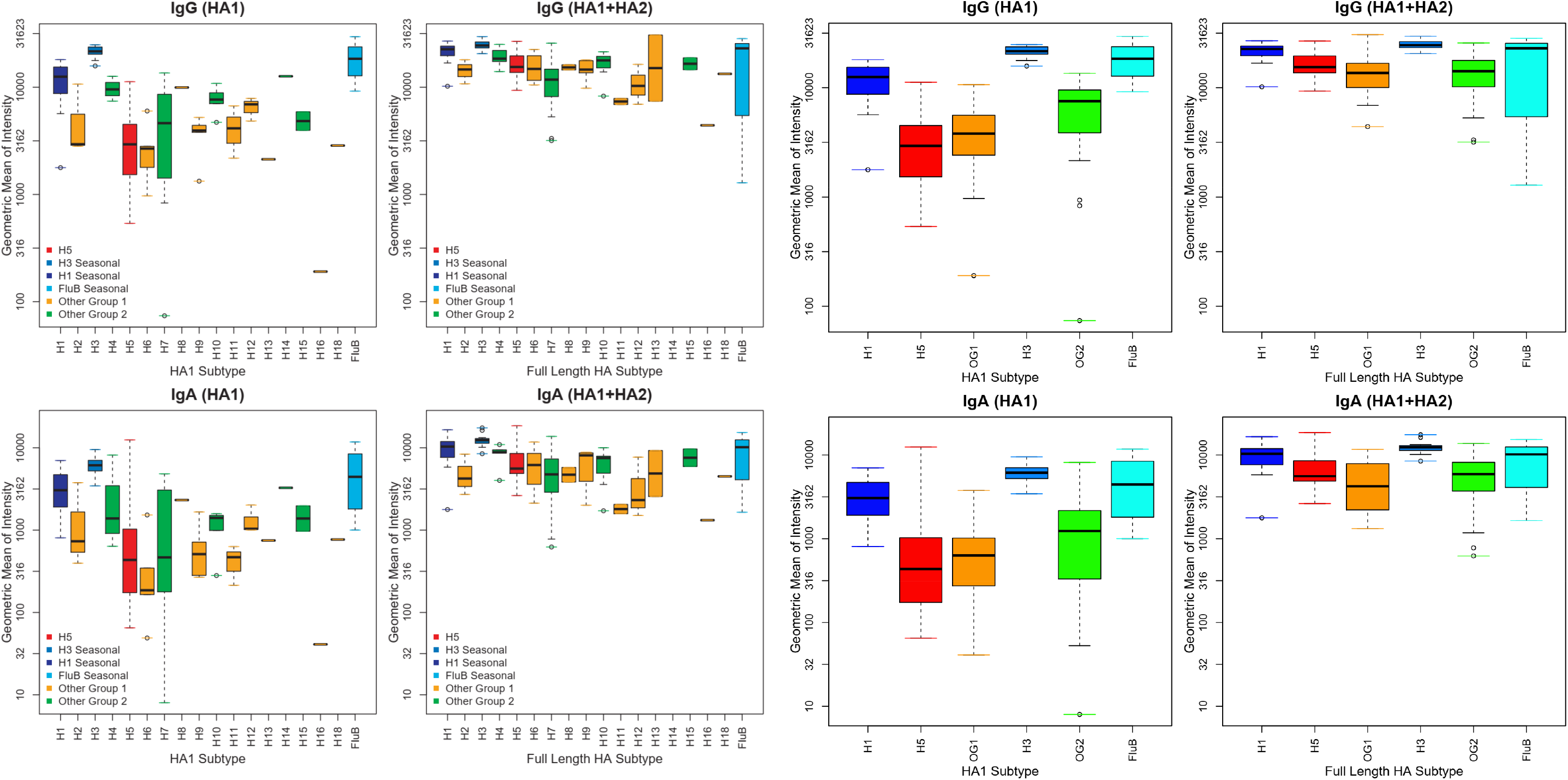
Pre-vaccination geometric mean intensity by HA subtype (Experiment 1). Each boxplot summarizes pre-vaccination geometric mean intensities by HA subtype across all 130 subjects. Results within subjects for multiple antigens of the same HA subtype were aggregated using the geometric mean.

**Supplemental Figure 1B.**
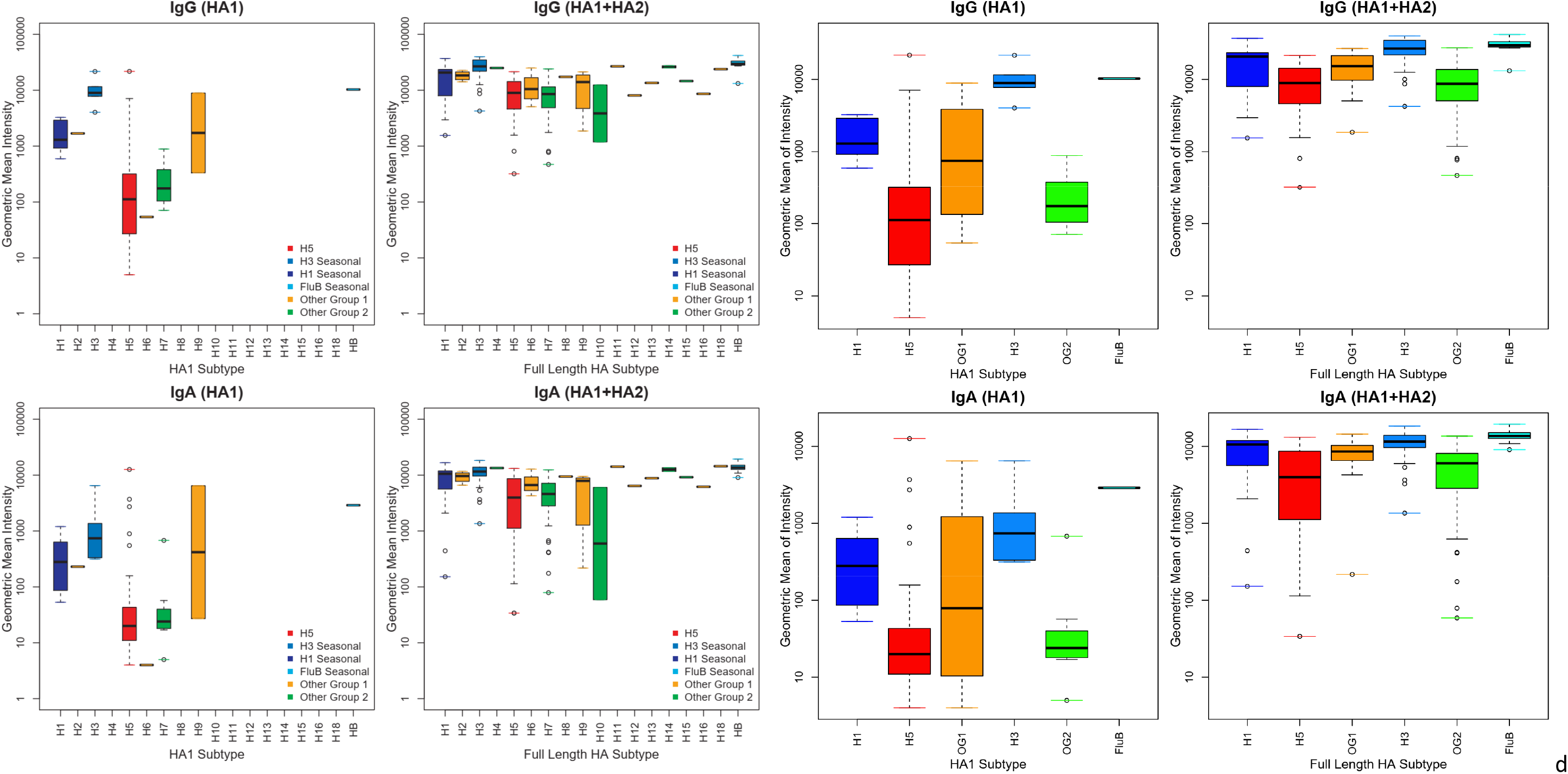
Pre-vaccination geometric mean intensity by HA subtype (Experiment 2). Each boxplot summarizes pre-vaccination geometric mean intensities by HA subtype across all 130 subjects. Results within subjects for multiple antigens of the same HA subtype were aggregated using the geometric mean.

**Supplemental Figure 2A.**
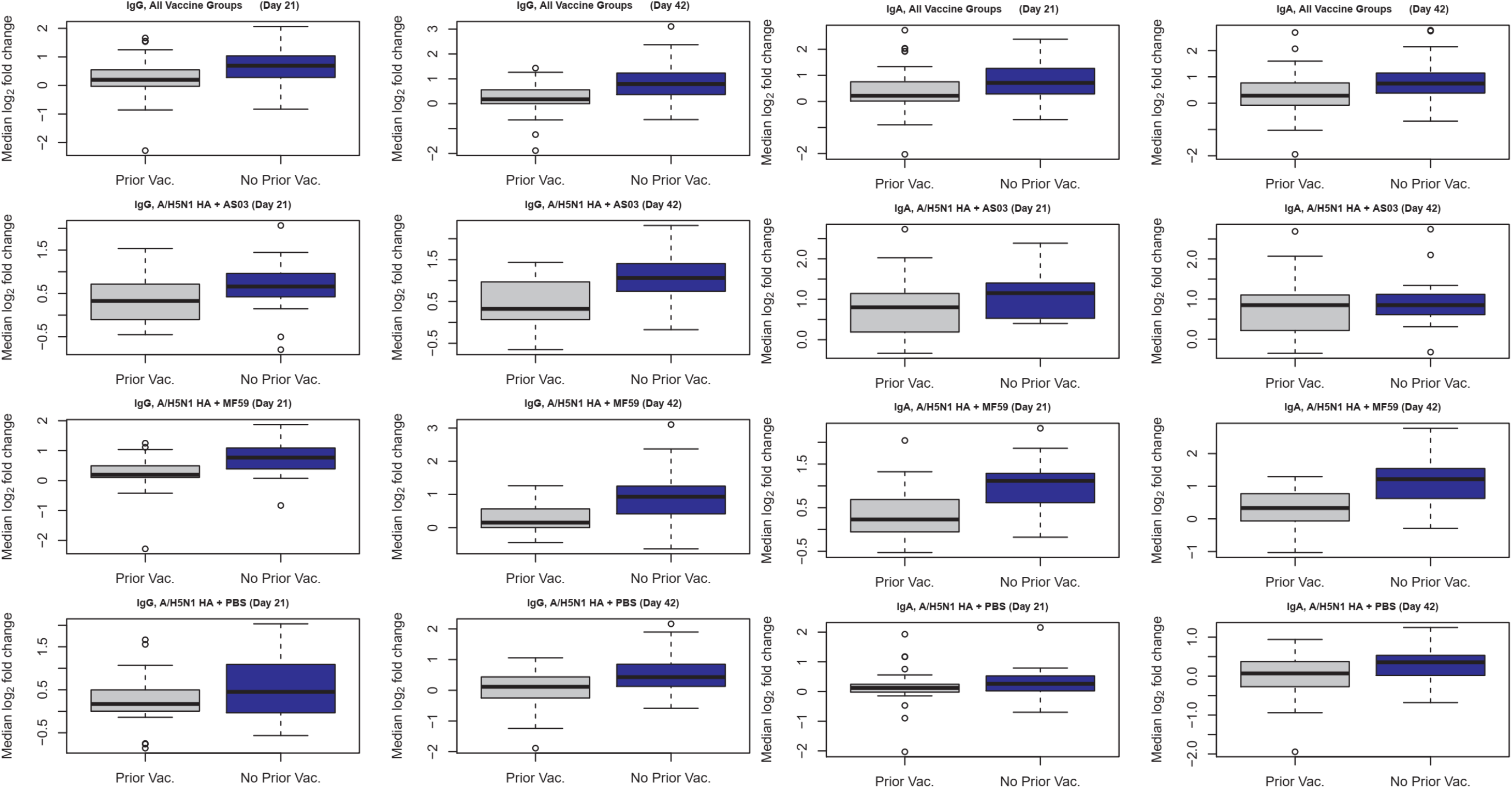
Impact of prior seasonal influenza vaccination on H5N1 vaccination effects (Experiment 1). Each boxplot summarizes the median log_2_ fold change for each post-vaccination day by vaccination status (subject receiving seasonal prior vaccinations within the past 2 years or those not receiving seasonal influenza vaccine within that time frame) by vaccine group. Results within subjects for multiple antigens were aggregated using the log_2_ median fold change.

**Supplemental Figure 2B.**
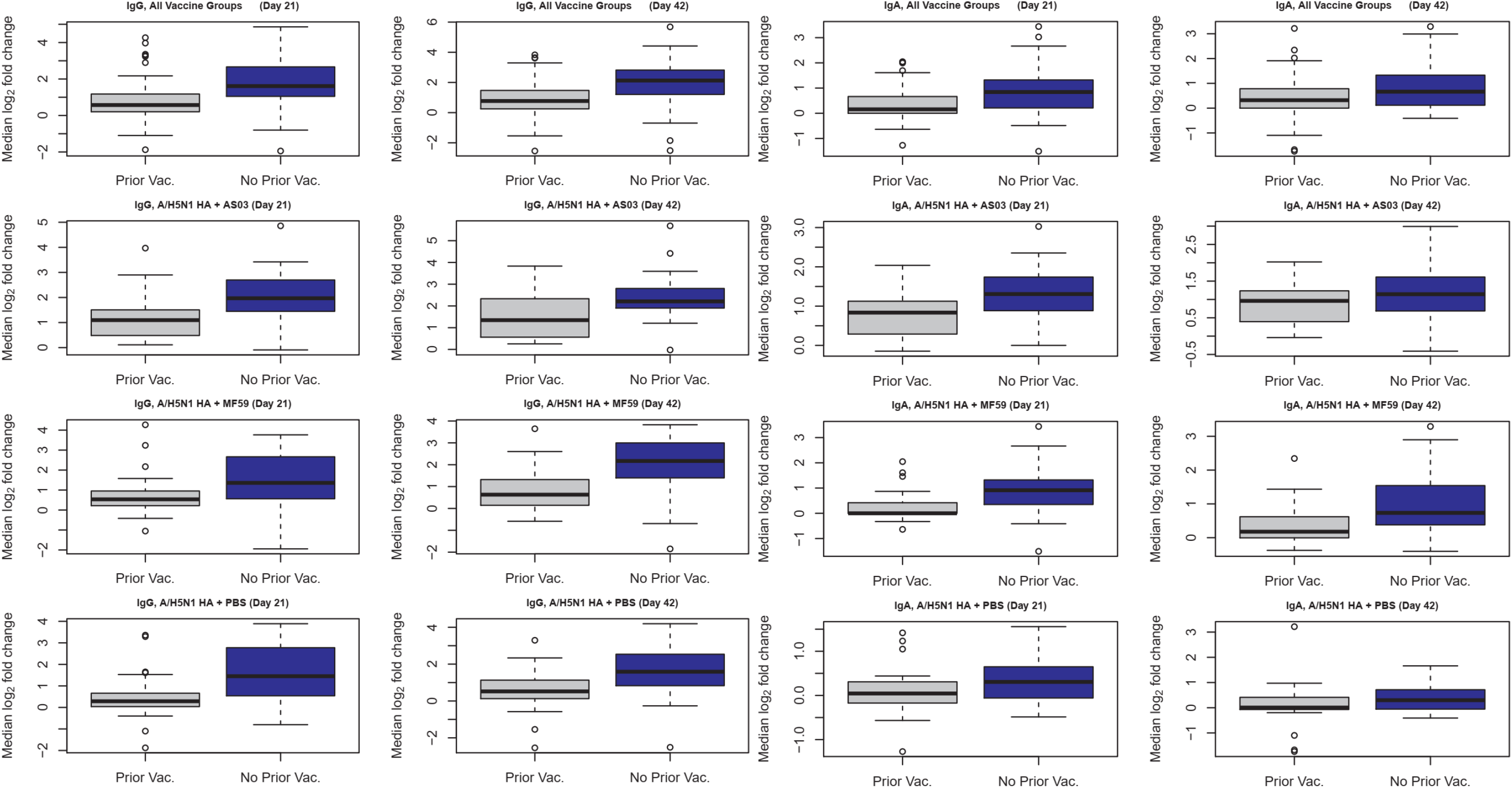
Impact of prior seasonal influenza vaccination on H5N1 vaccination effects (Experiment 2). Each boxplot summarizes the median log_2_ fold change for each post-vaccination day by vaccination status (subject receiving seasonal prior vaccinations within the past 2 years or those not receiving seasonal influenza vaccine within that time frame) by vaccine group. Results within subjects for multiple antigens were aggregated using the log_2_ median fold change.

**Supplemental Figure 3.**
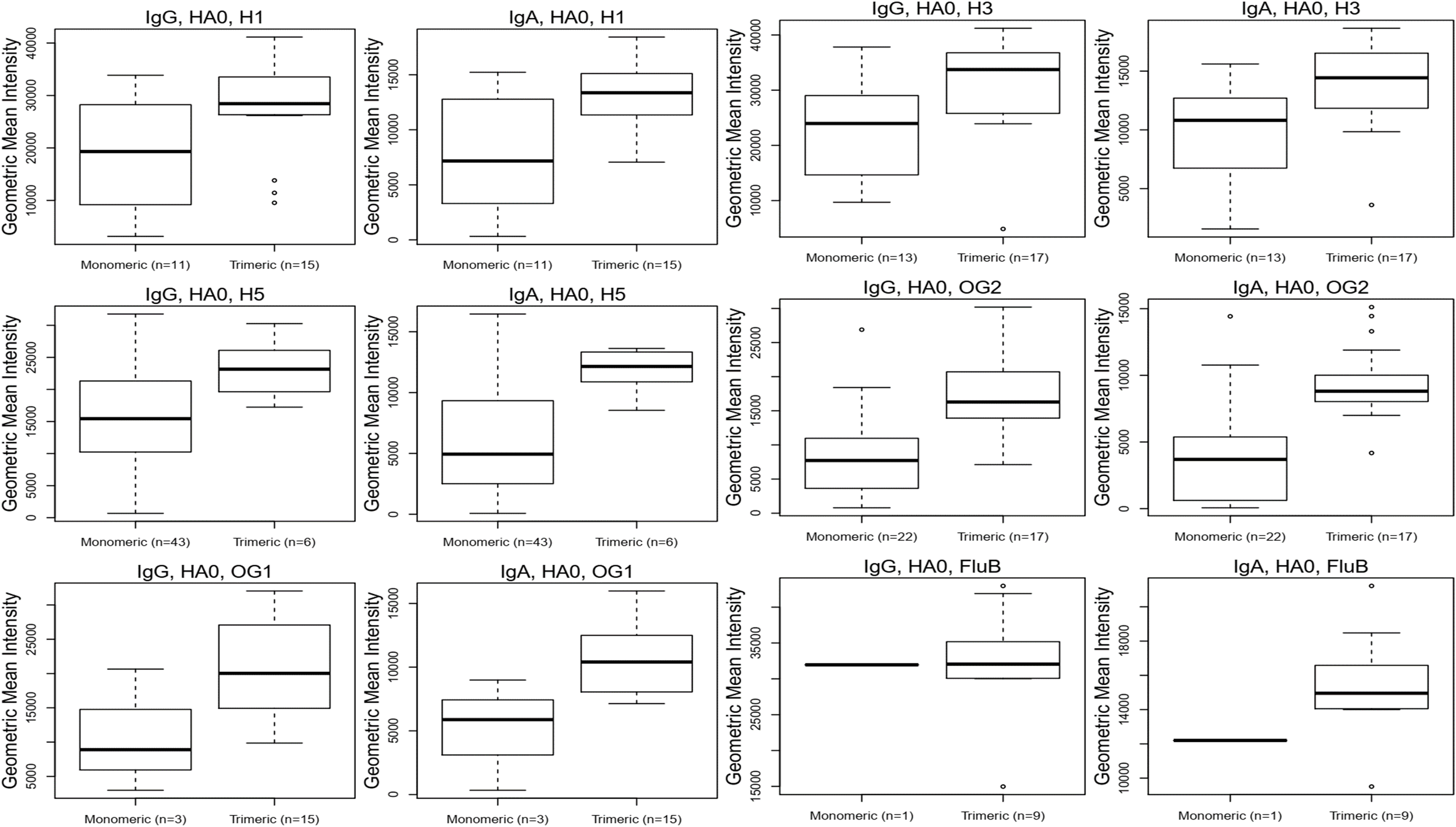
Comparison of trimeric vs. non-trimeric HA0 response by subtype (Experiment 2). Antibody reactivity of monomeric HA0s was compared with stabilized trimeric HA0s. Each boxplot summarizes the geometric mean intensity across all samples by antigen type (trimeric vs. monomeric) irrespective of post-vaccination day and vaccine group. Results within subjects for multiple antigens were aggregated using the geometric mean intensity.

**Supplemental Figure 4.**
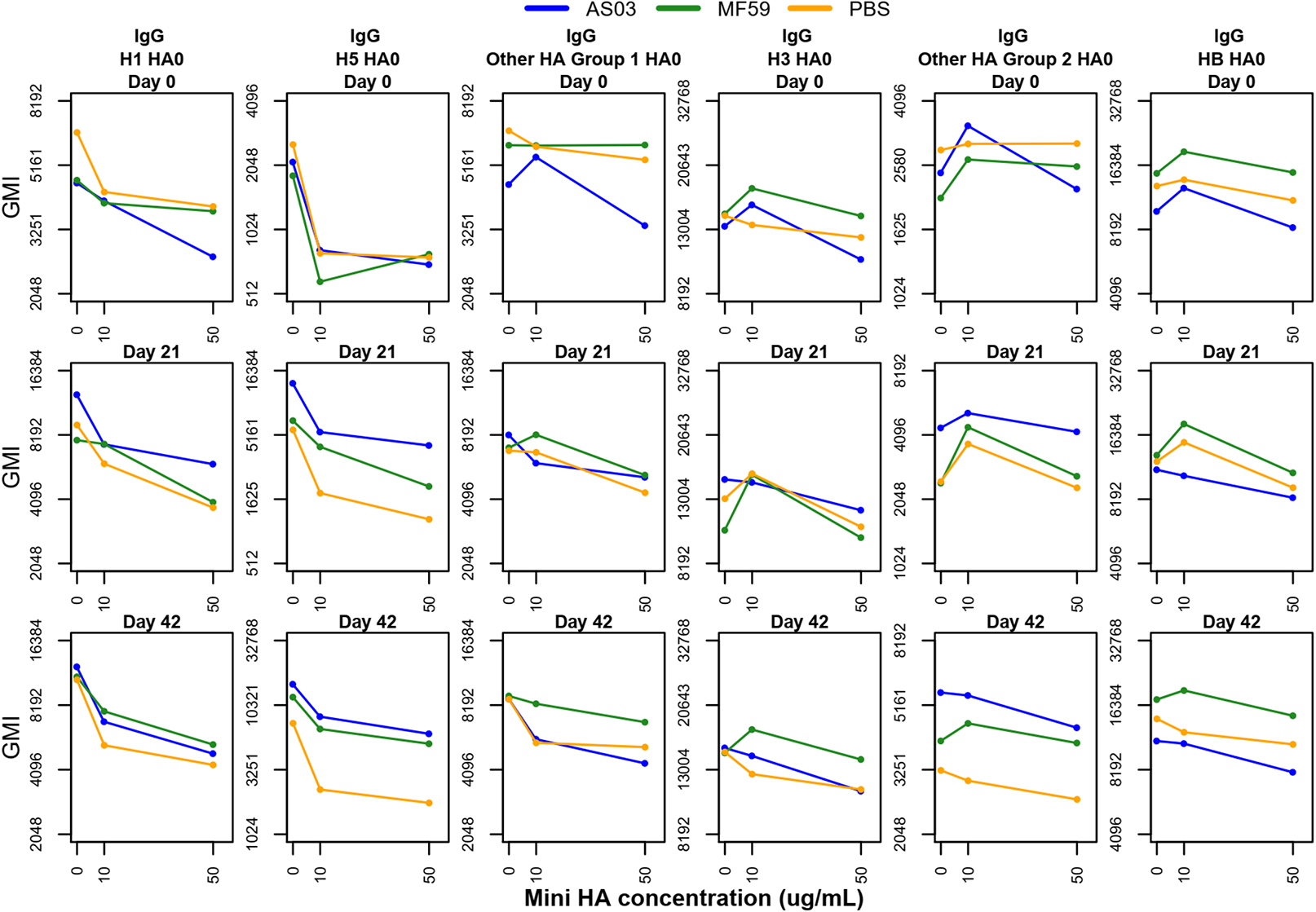
IgG anti-stalk blocking effect by mini-HA concentration and HA subtype. Each trend line summarizes the geometric mean intensity by vaccine group, HA subtype across increasing mini-HA concentrations. Results within subjects for multiple antigens were aggregated using the geometric mean intensity.

**Supplemental Figure 5.**
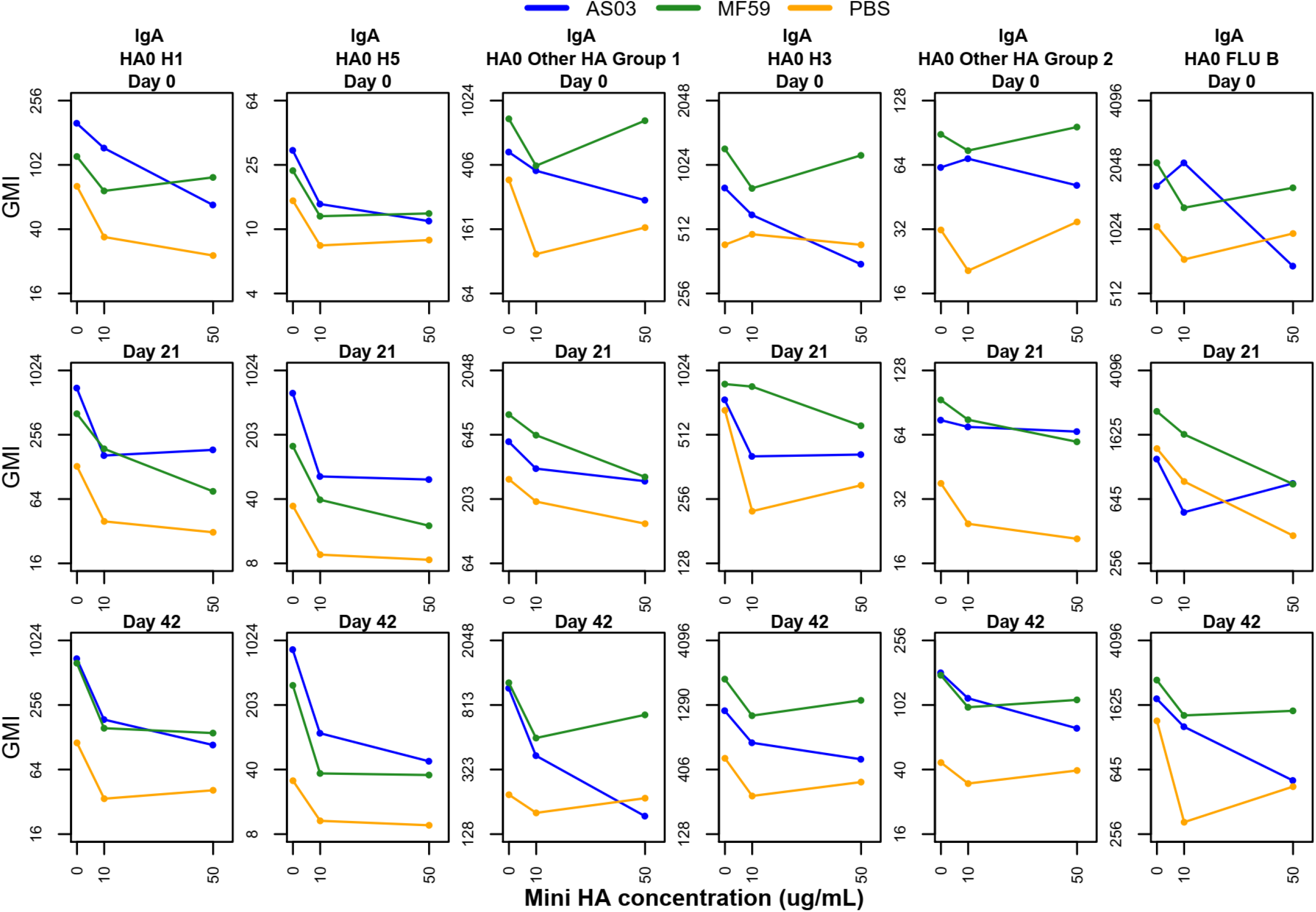
IgA anti-stalk blocking effect by mini-HA concentration and HA subtype. Each trend line summarizes the geometric mean intensity by vaccine group, HA subtype across increasing mini-HA concentrations. Results within subjects for multiple antigens were aggregated using the geometric mean intensity.

**Supplemental Figure 6.**
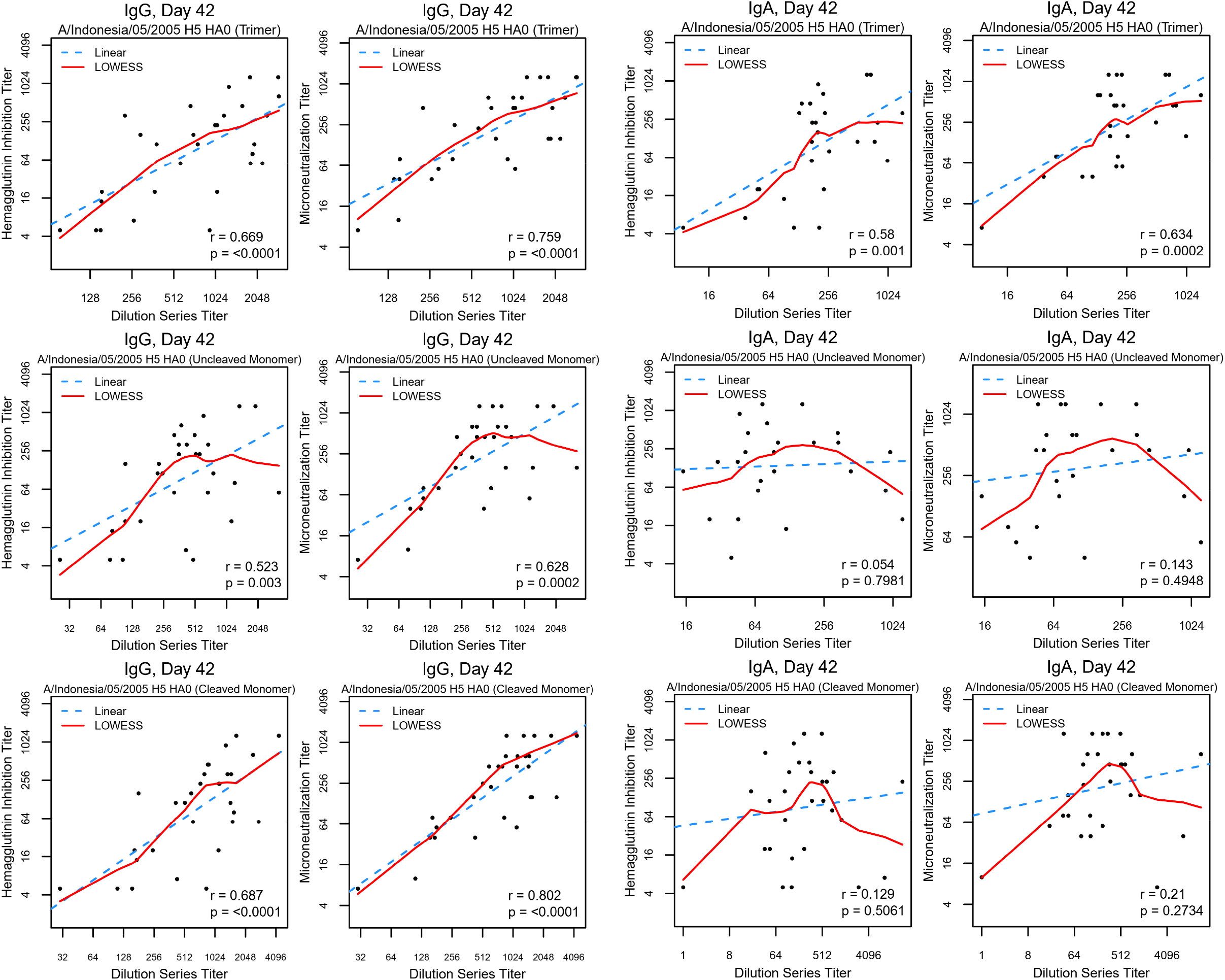
Associations between dilution-based titers based on microarray results and HAI and Microneutralization Titer. Results for 4 vaccine antigens are shown. The red line represents a locally weighted regression fit while the blue dashed line indicates a linear fit.

## SUPPLEMENTAL TABLES

**Supplemental Table 1A.** Demographics (Sex and Previous Vaccination)

|  |  | A/H5N1 HA +<br>AS03 (N=38) |  | A/H5N1 HA +<br>MF59 (N=42) |  | A/H5N1 HA +<br>PBS (N=50) |  |
| --- | --- | --- | --- | --- | --- | --- | --- |
|  |  | N | % | N | % | N | % |
| Sex | Female | 15 | 39.50% | 13 | 31% | 18 | 36% |
|  | Male | 23 | 60.50% | 29 | 69% | 32 | 64% |
| Previous<br>vaccination | No | 19 | 50% | 18 | 42.90% | 20 | 40% |
|  | Yes | 19 | 50% | 24 | 57.10% | 30 | 60% |

**Supplemental Table 1B.**
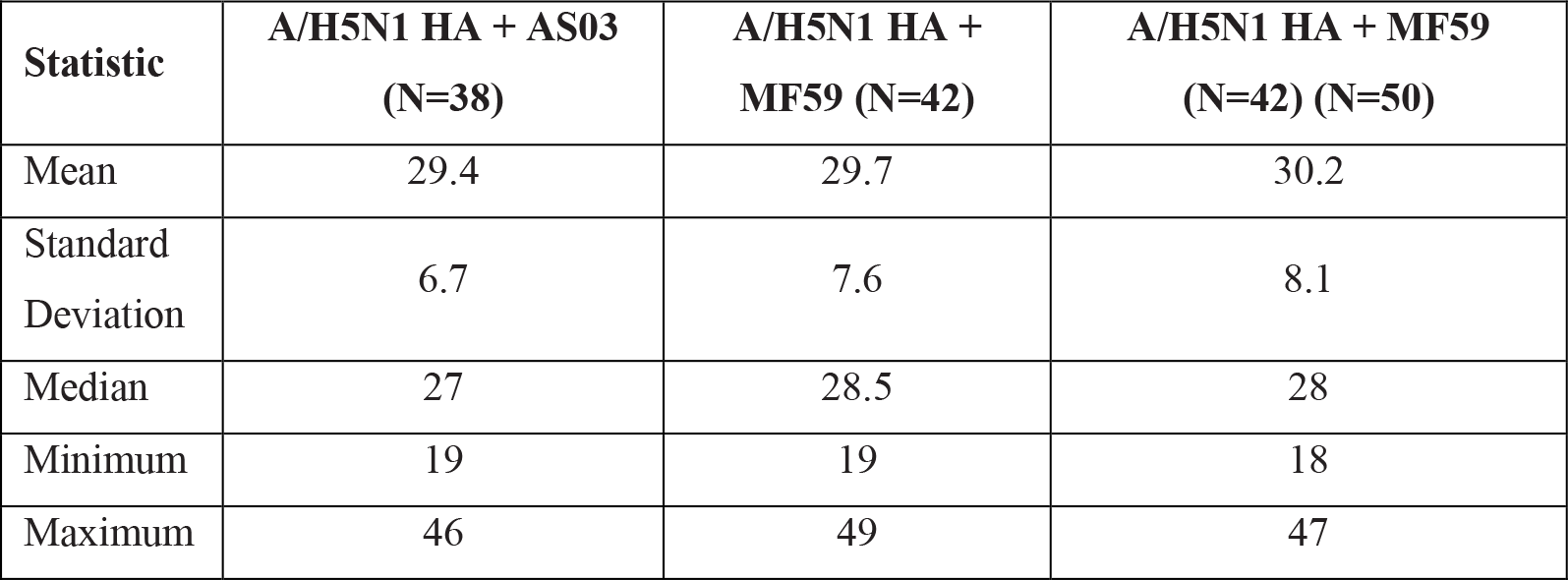
Demographics (Age Distribution)

**Supplemental Table 2.**
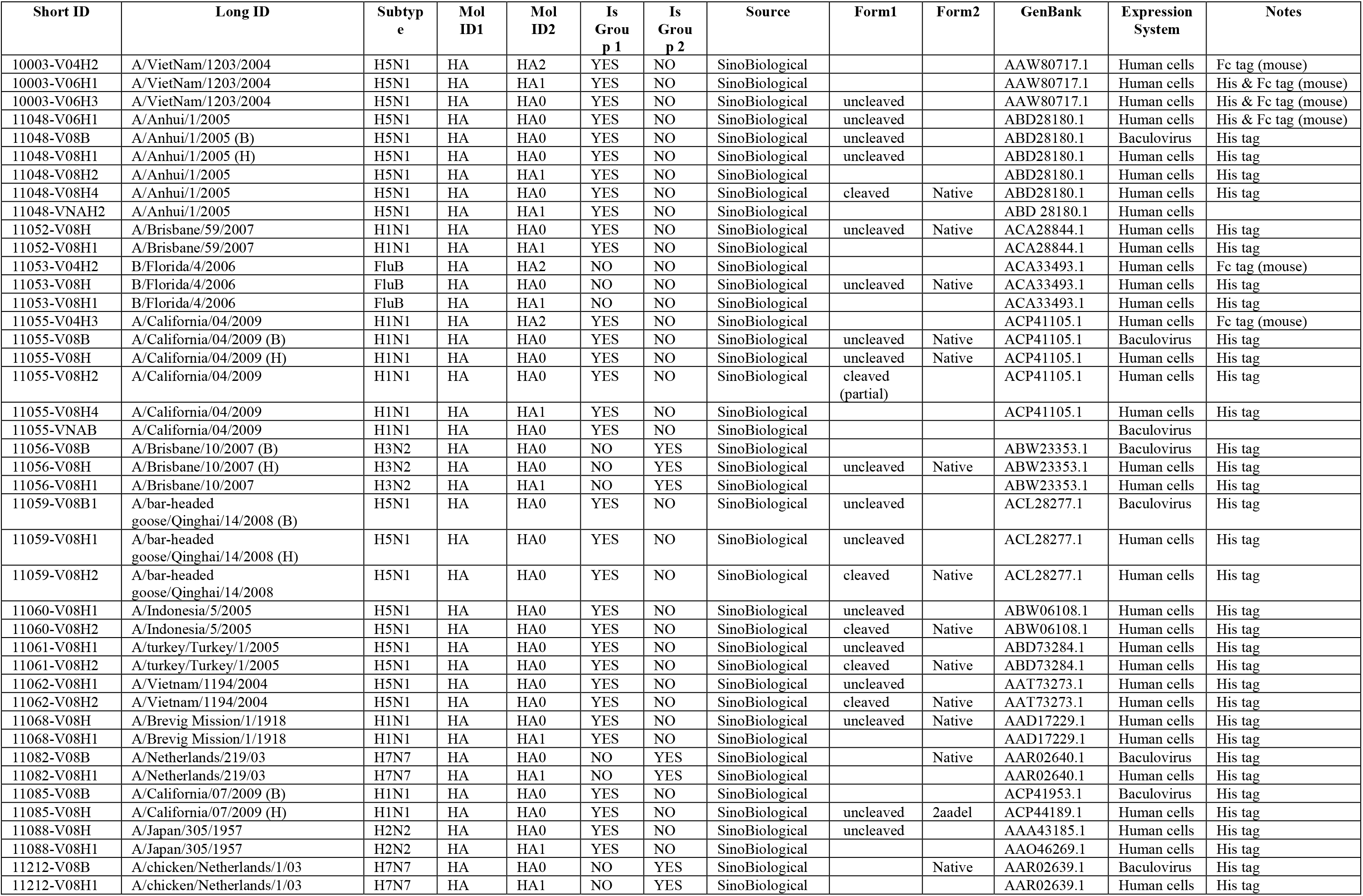
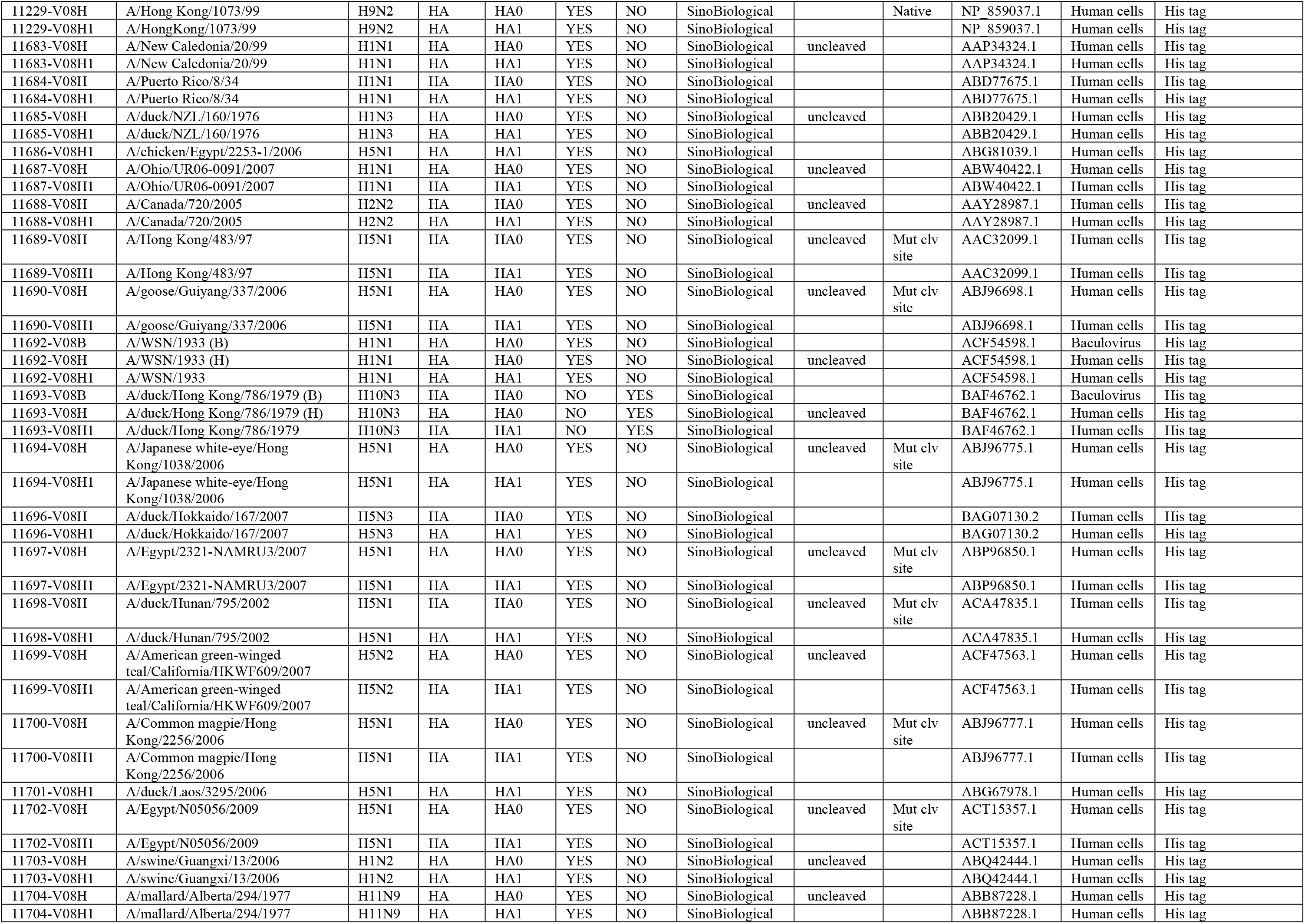
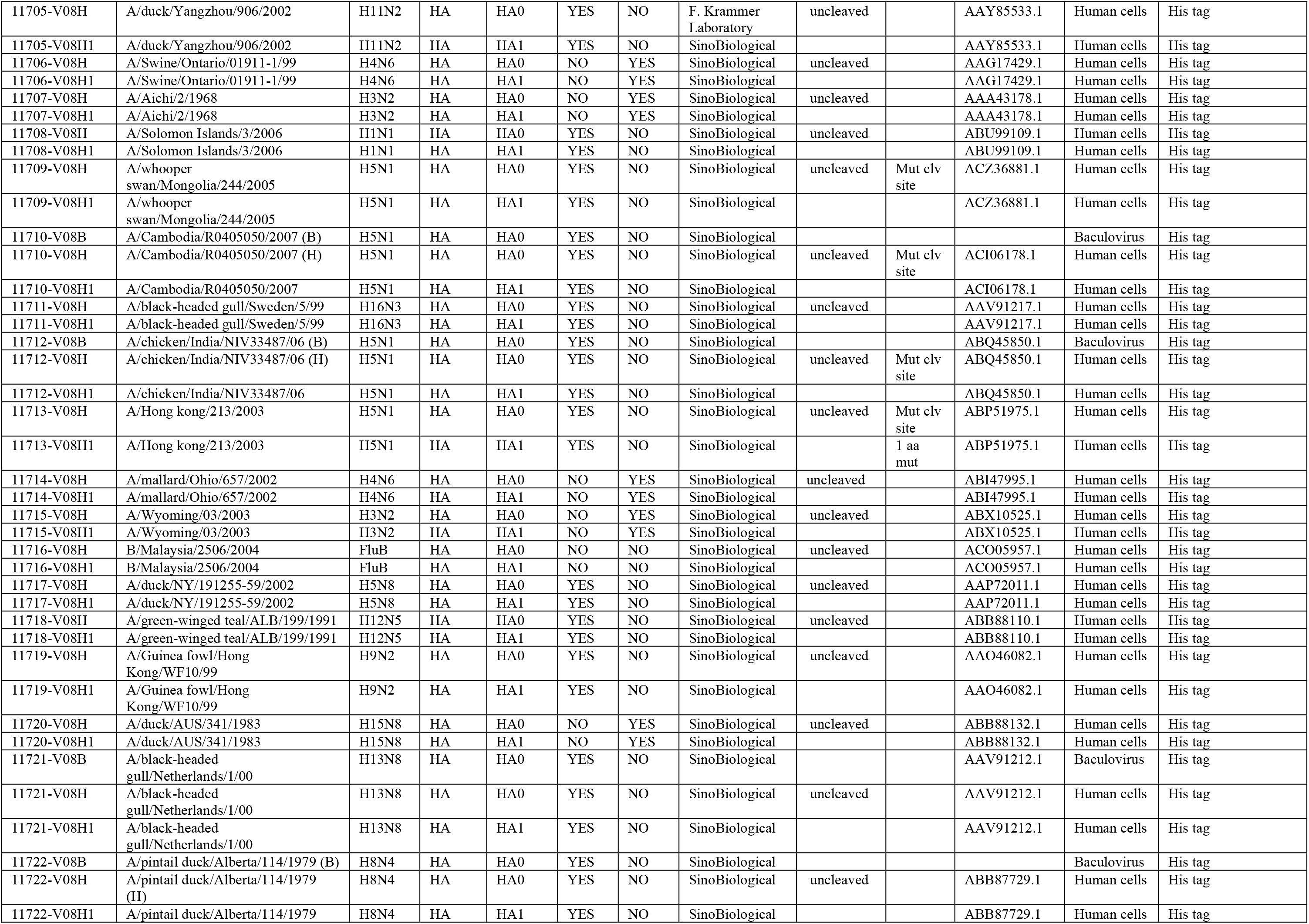
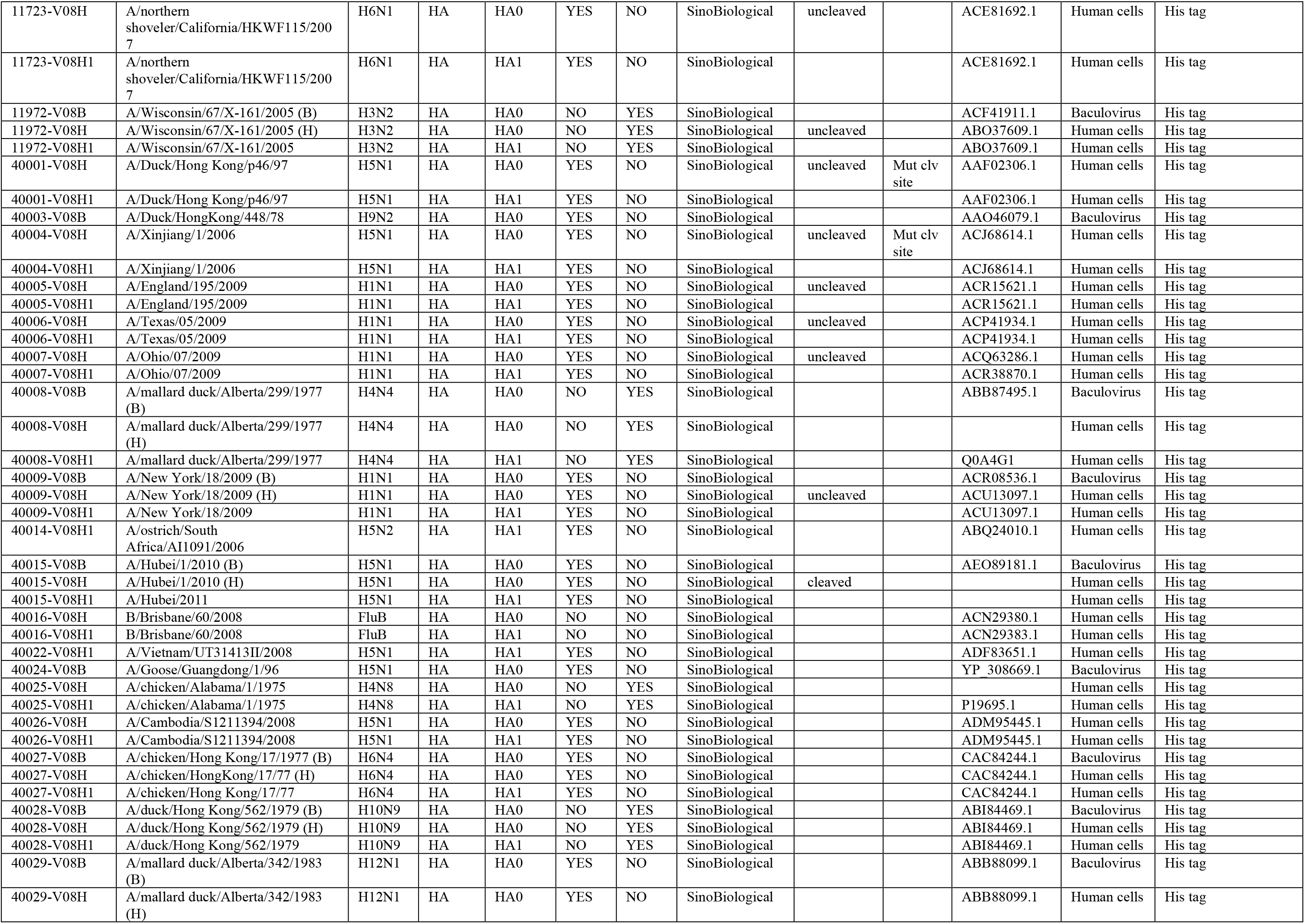
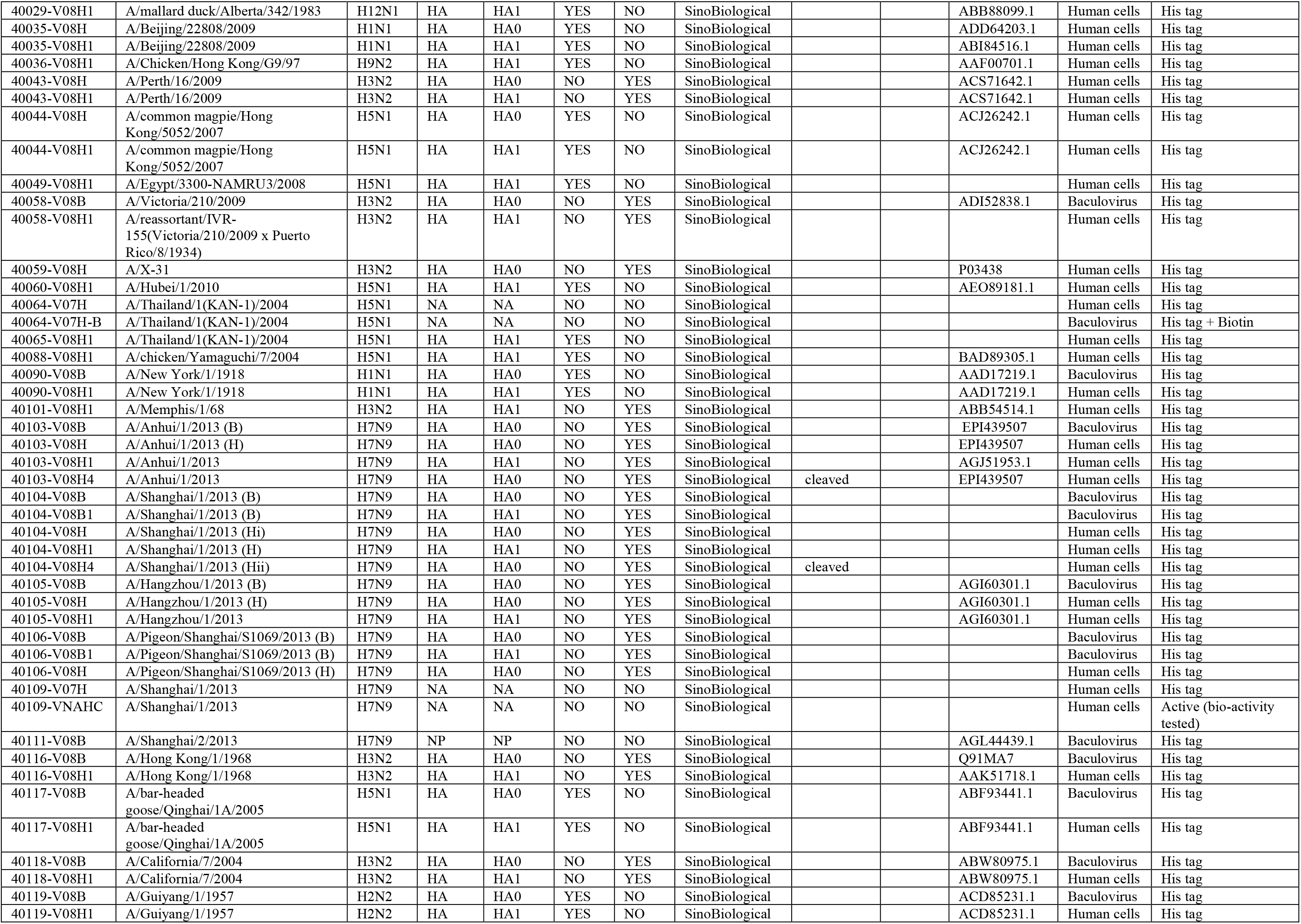
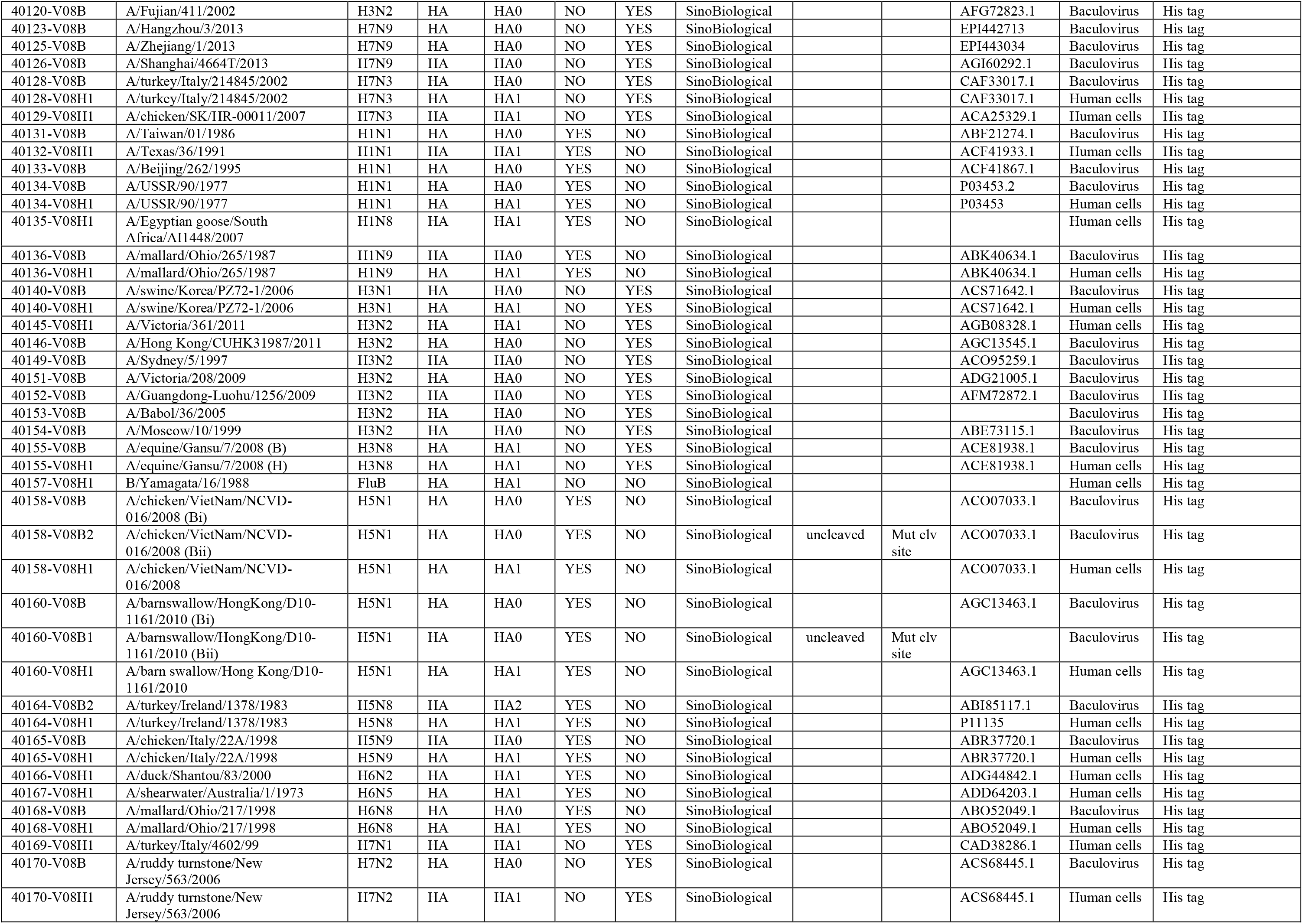
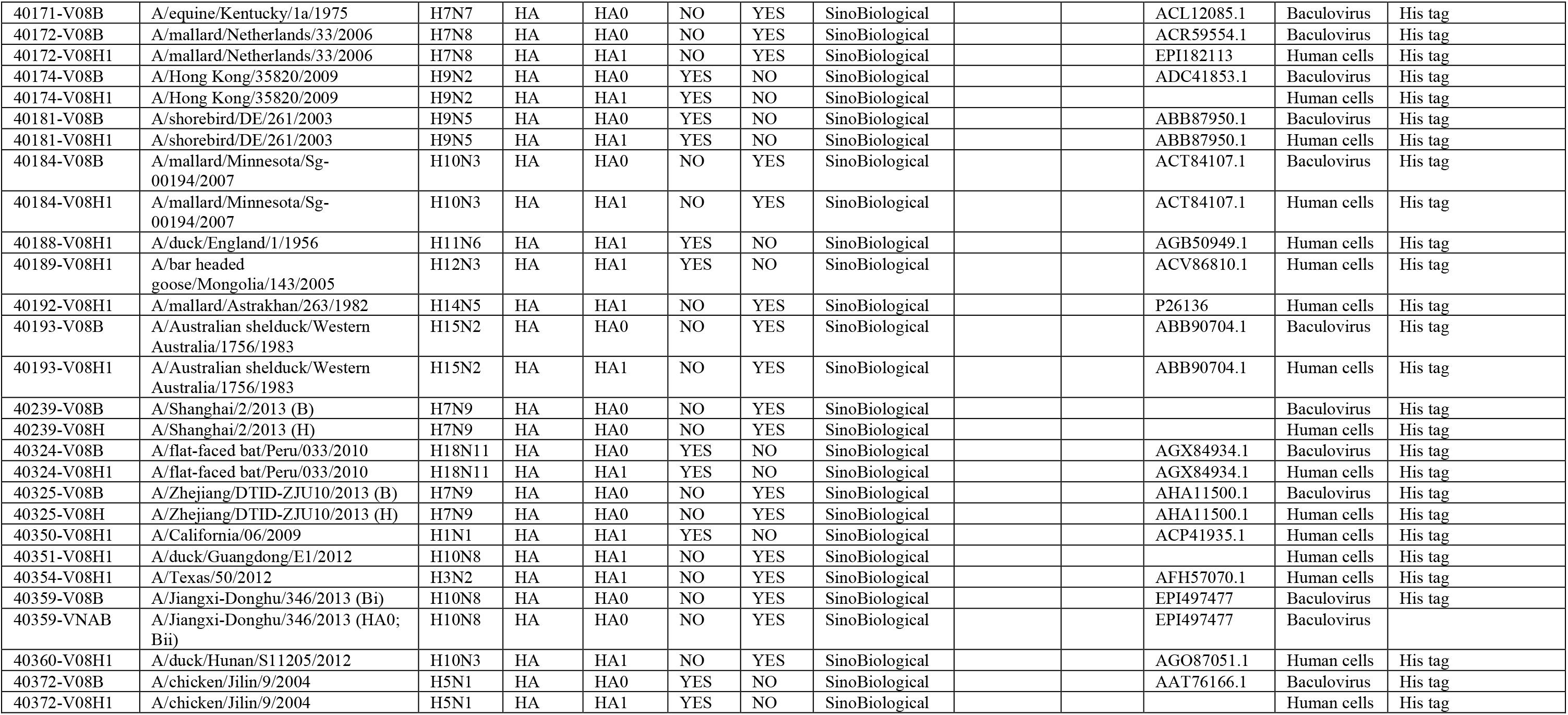
List of Proteins on Array #1.

**Supplemental Table 3.**
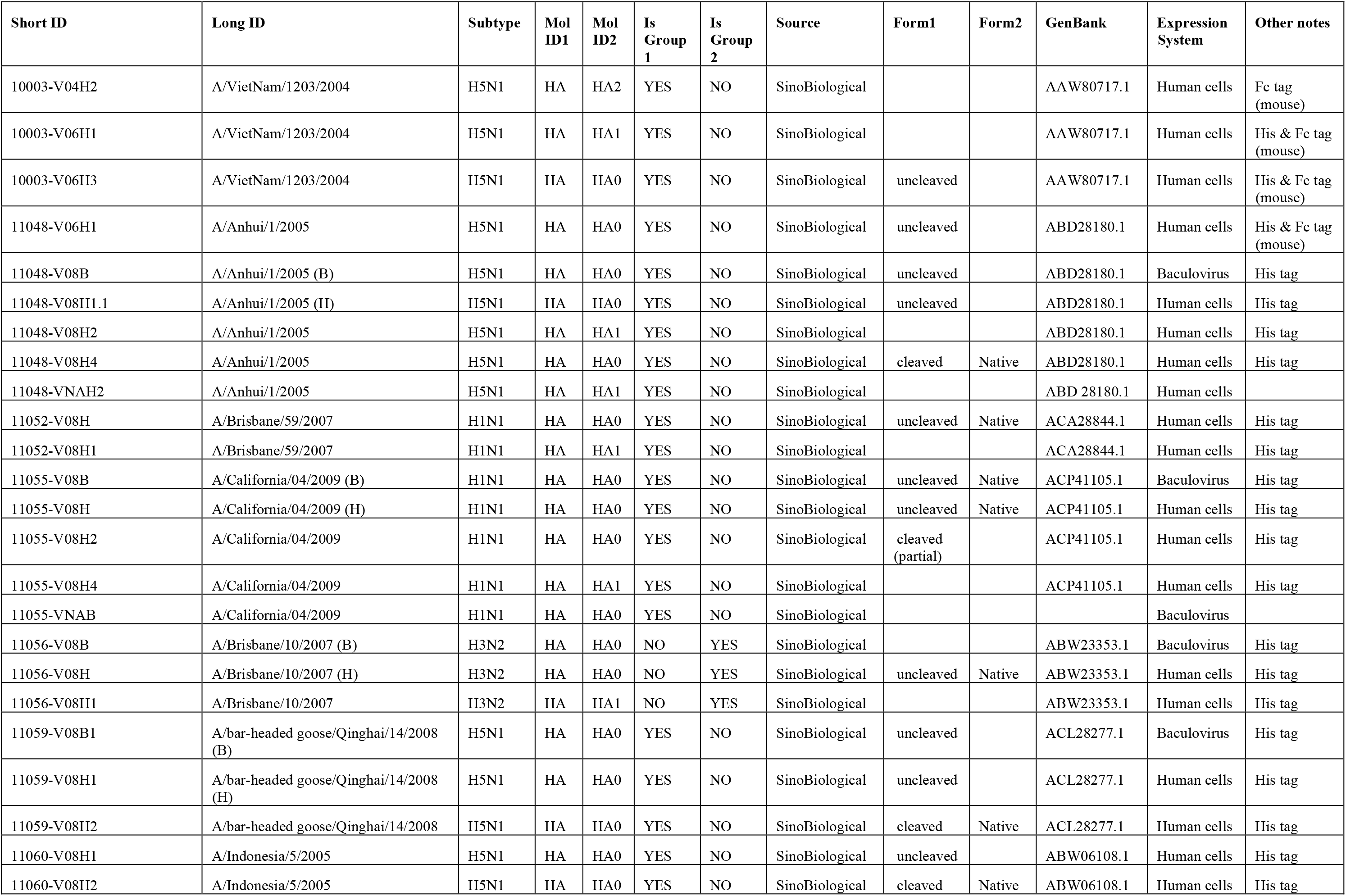
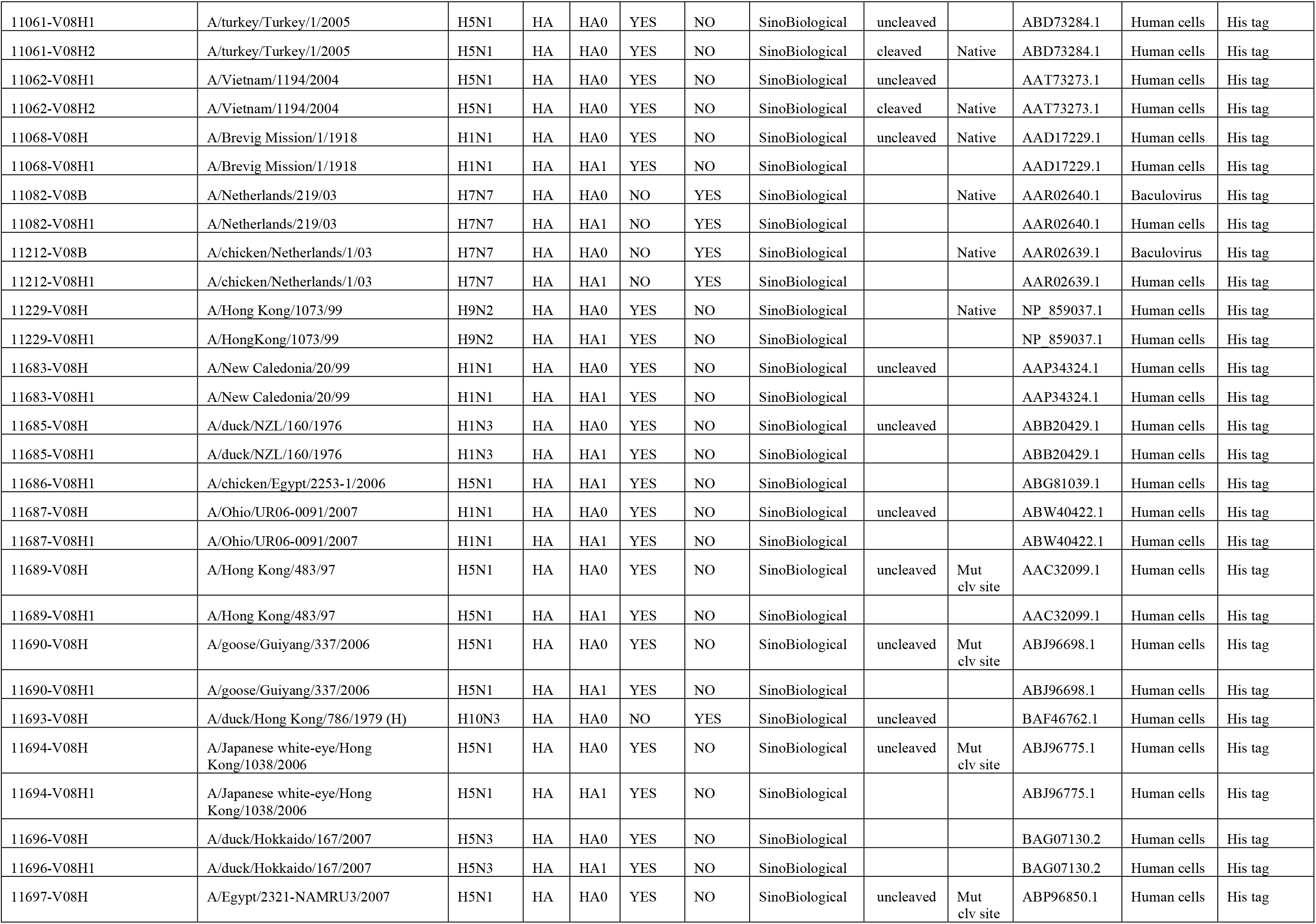
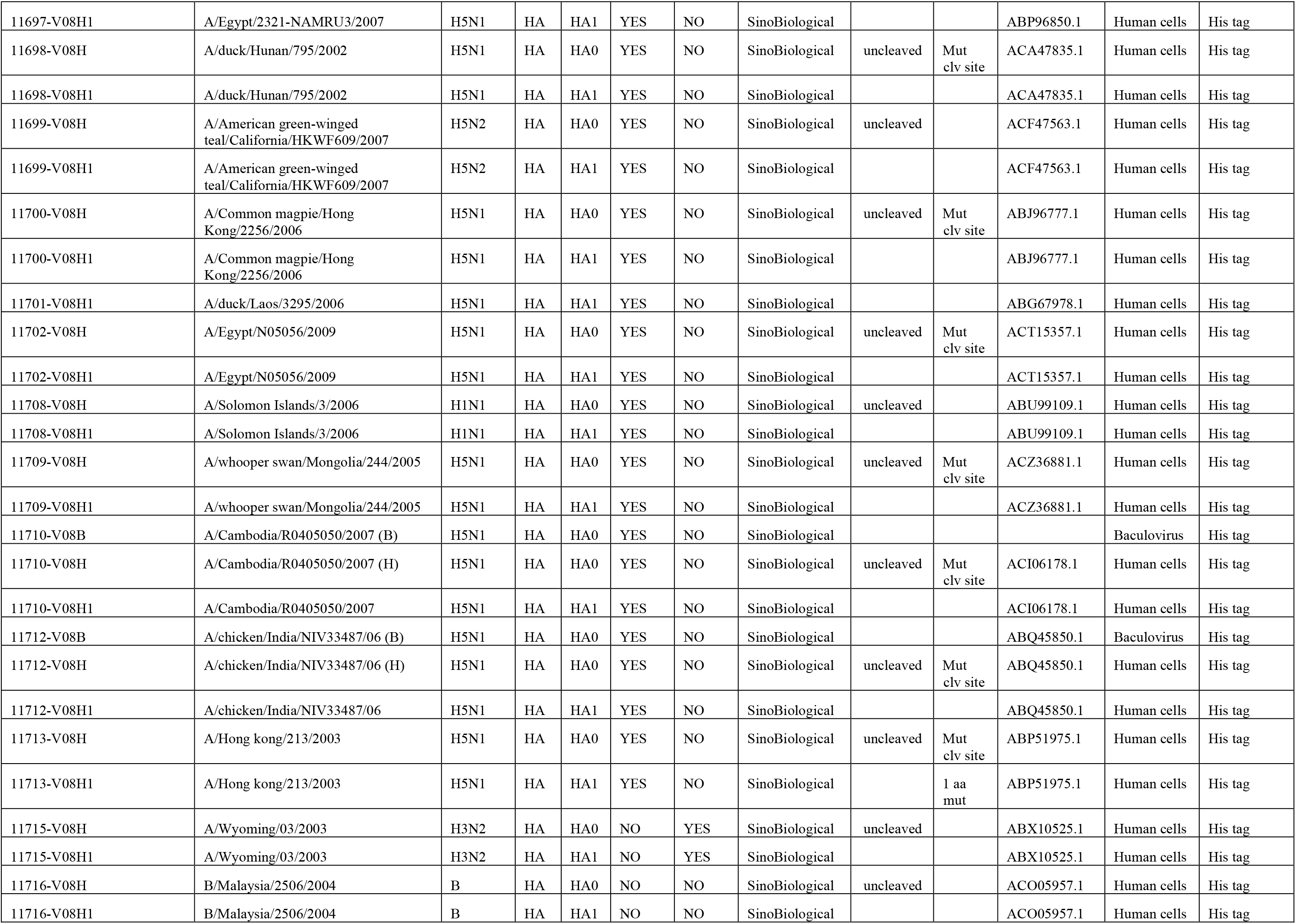
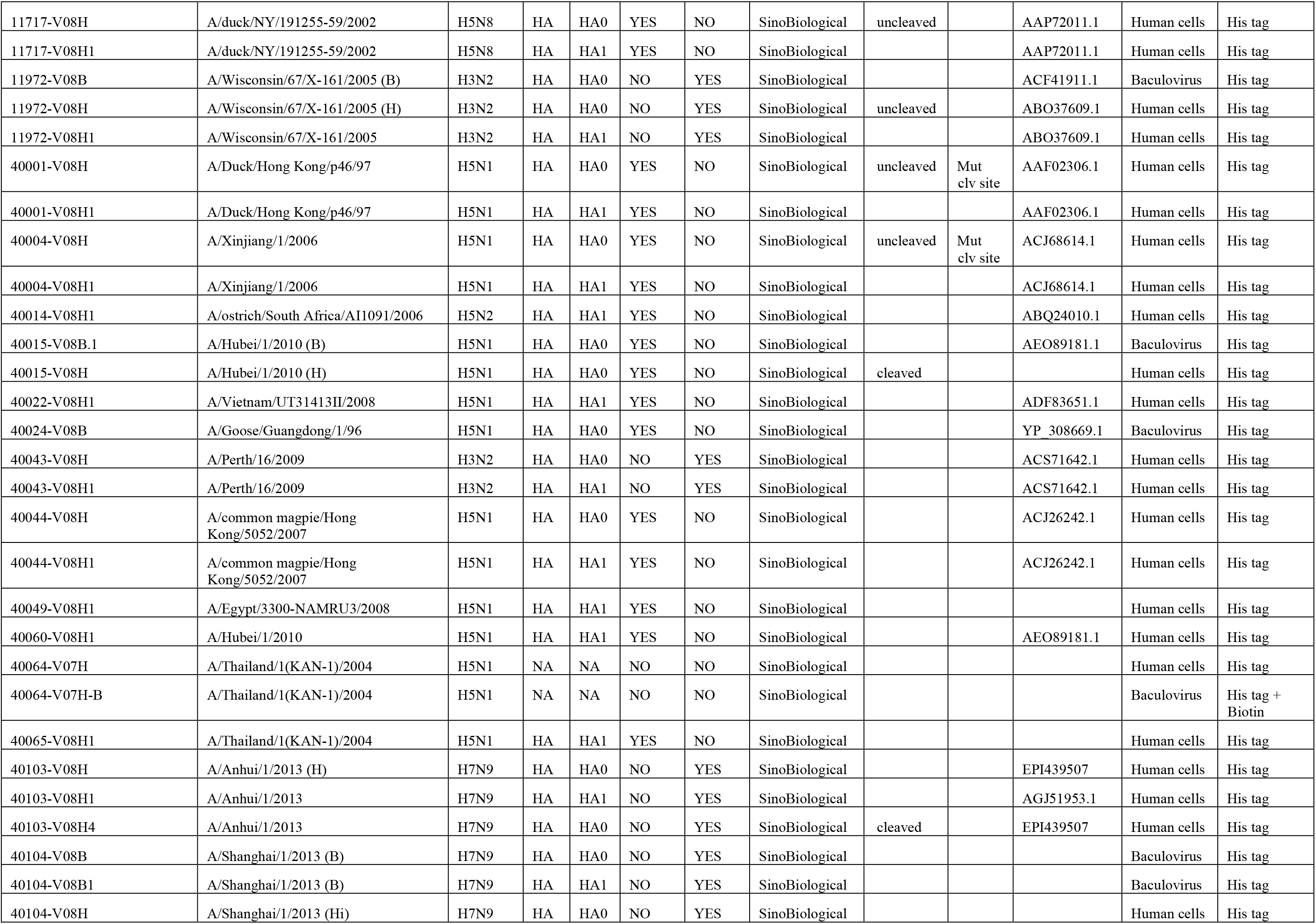
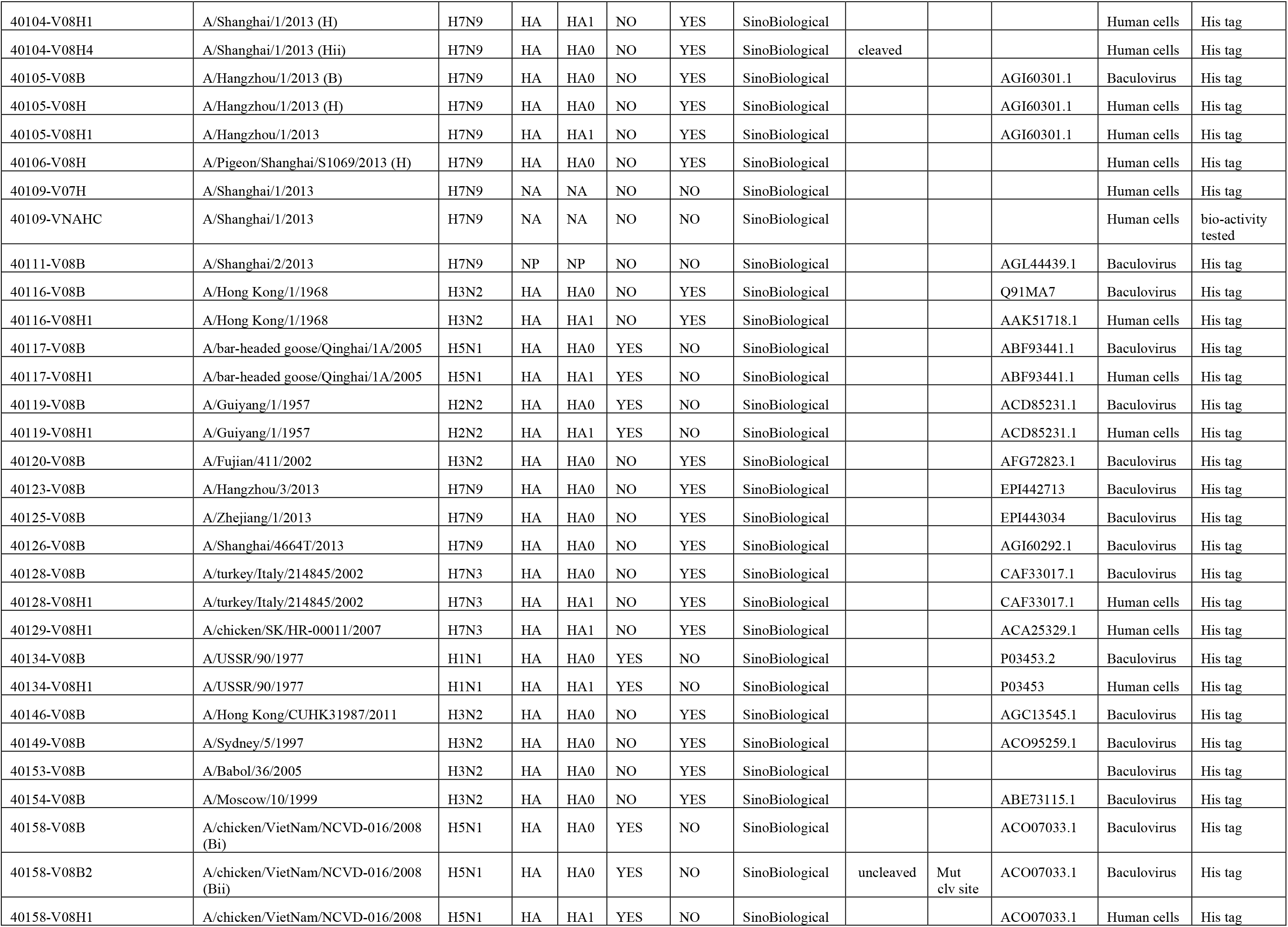
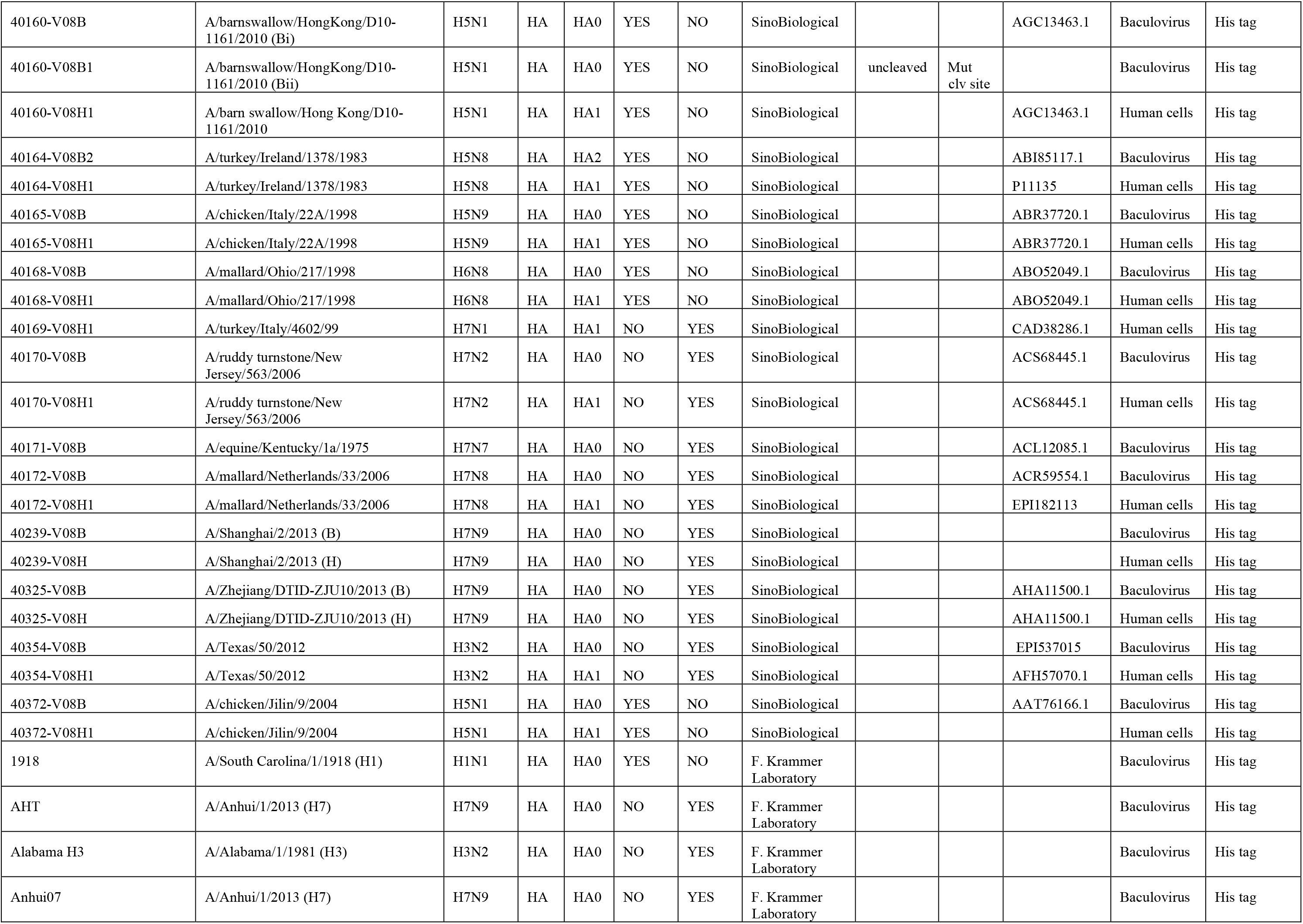
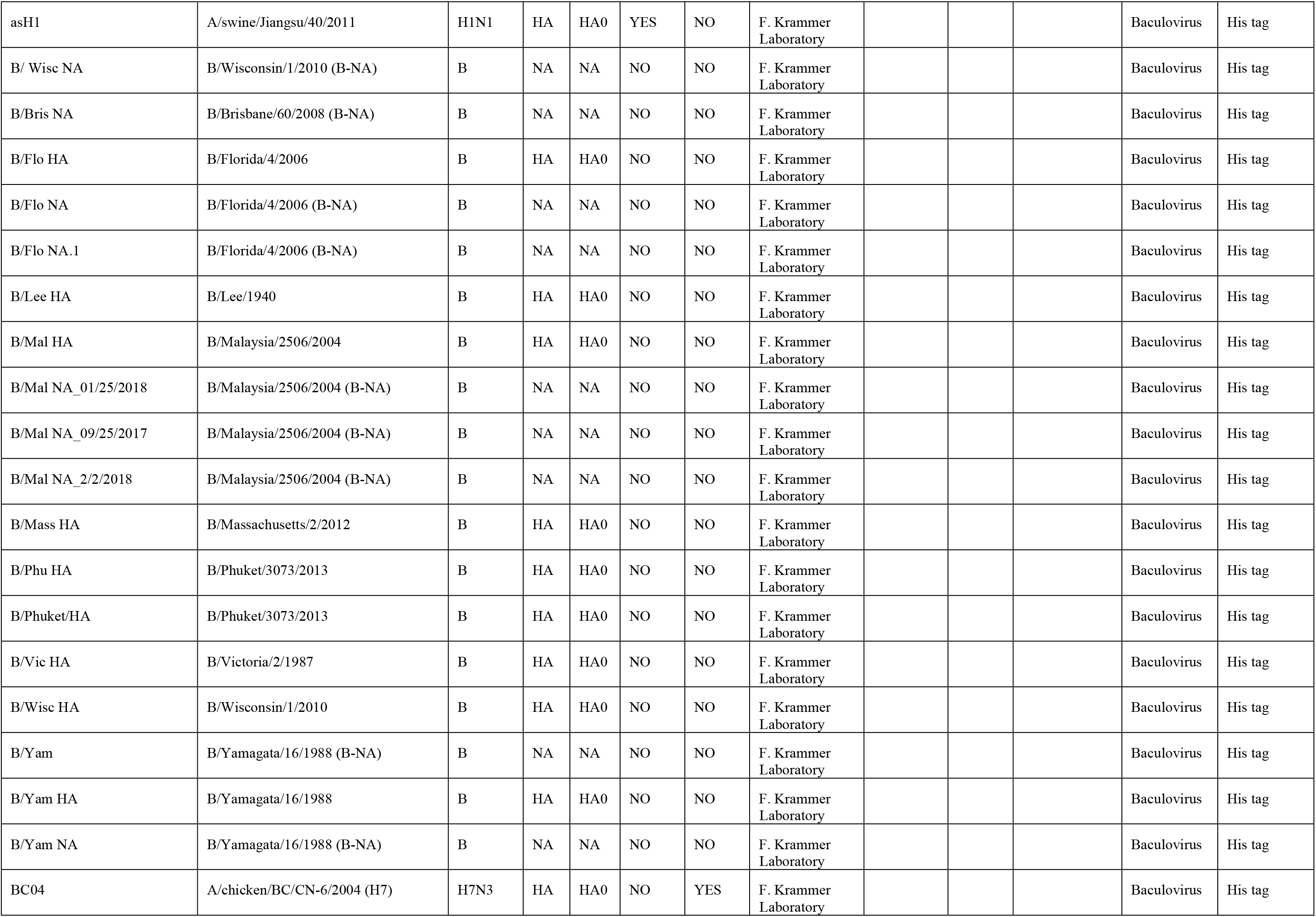
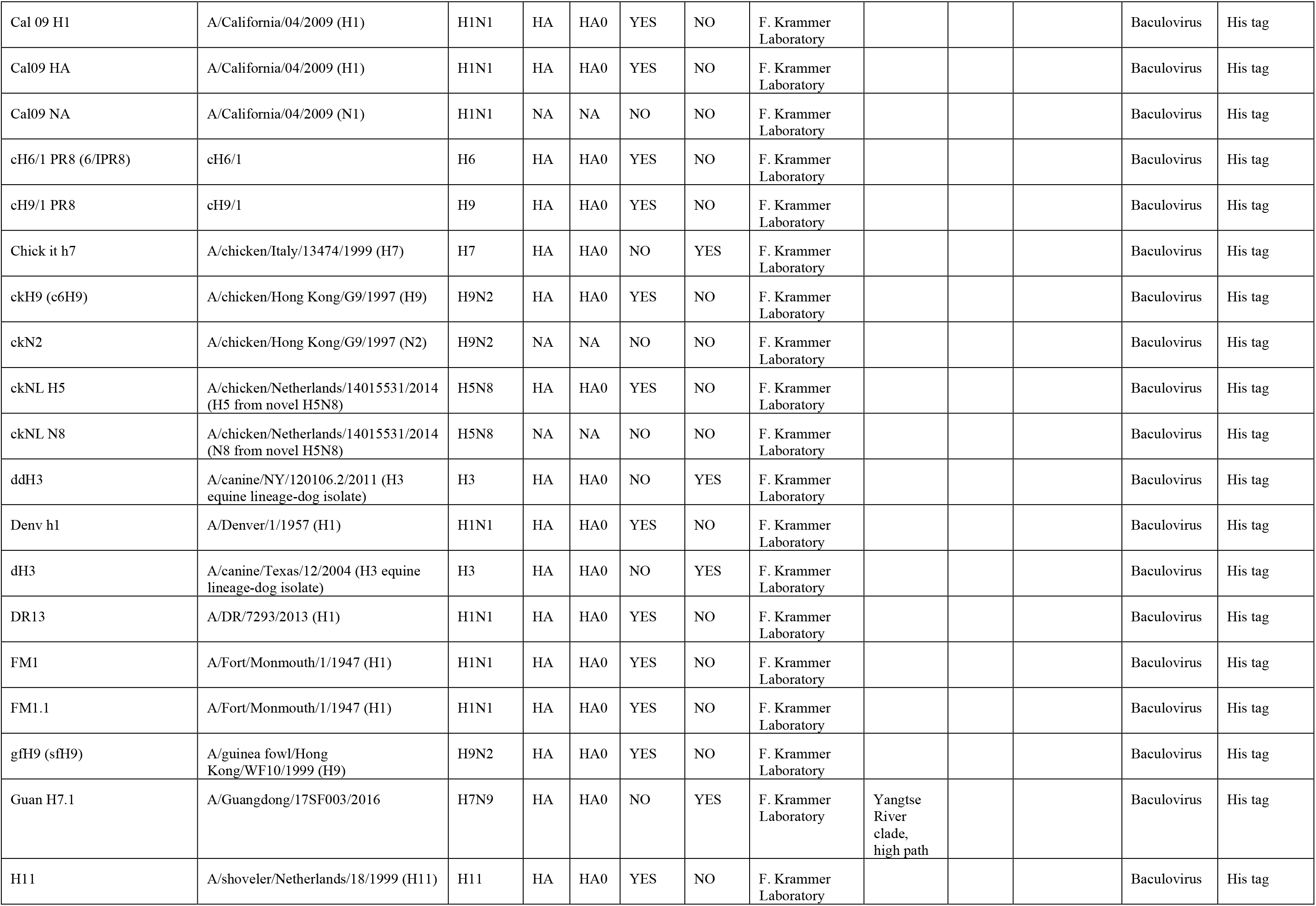
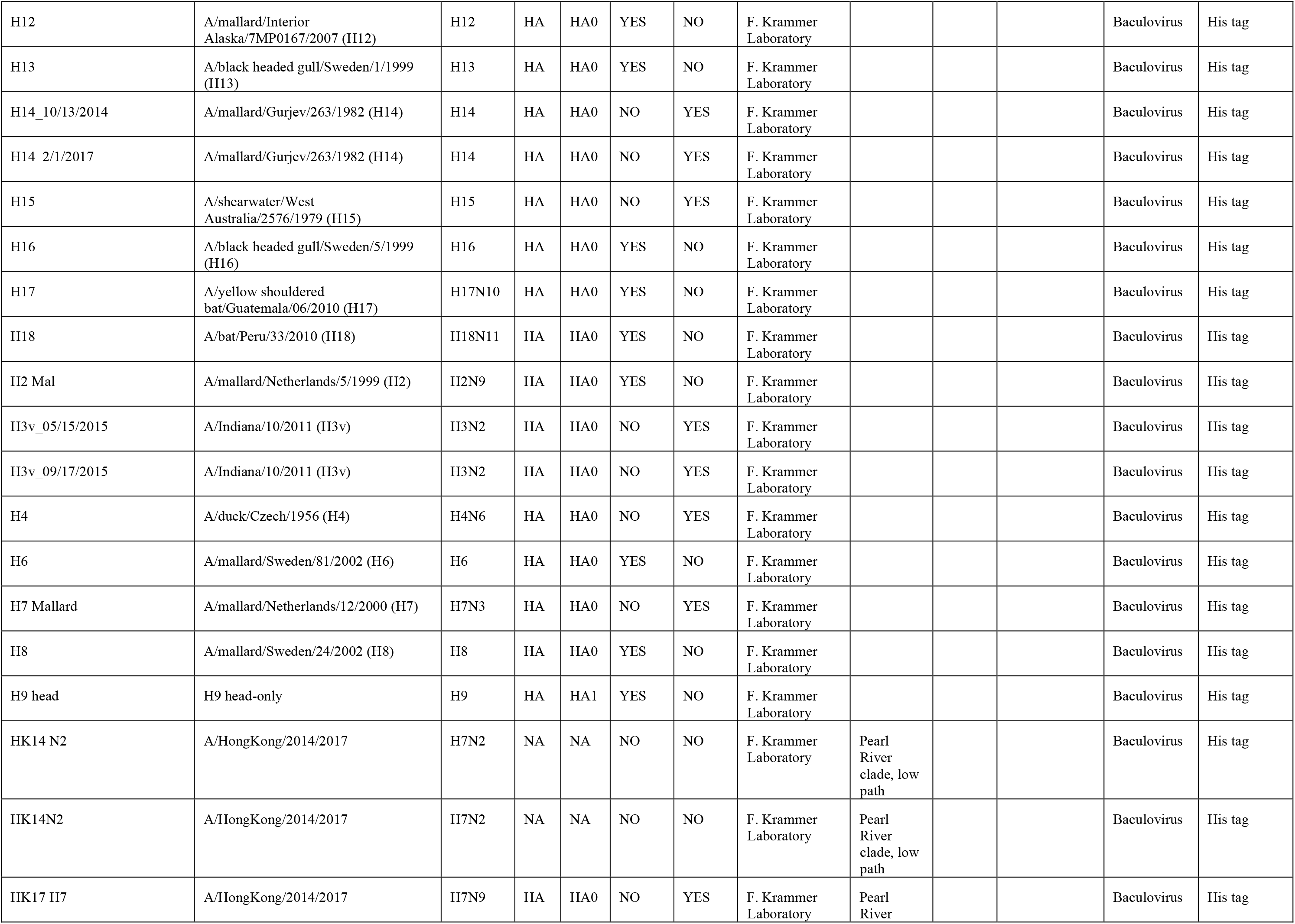
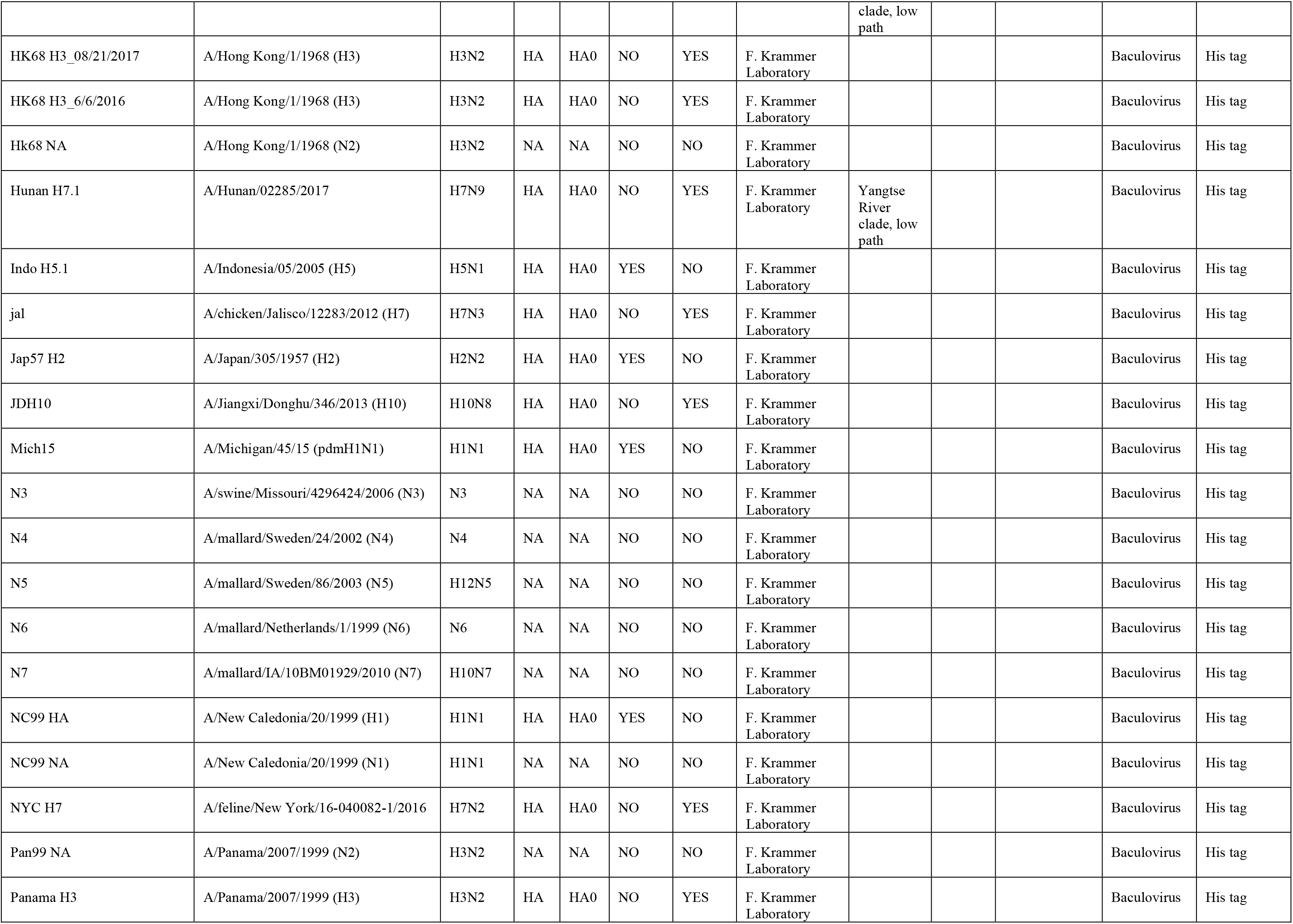
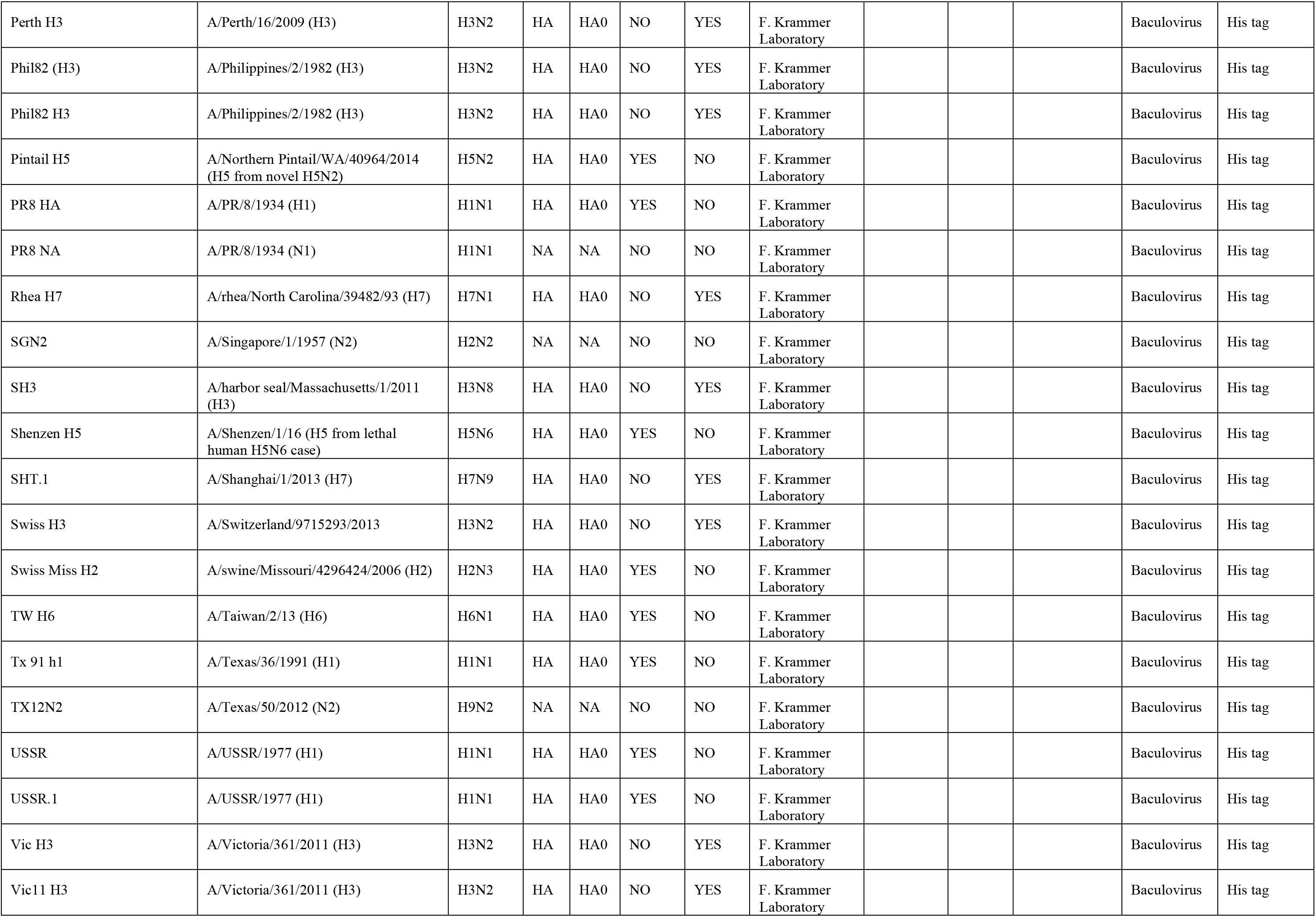
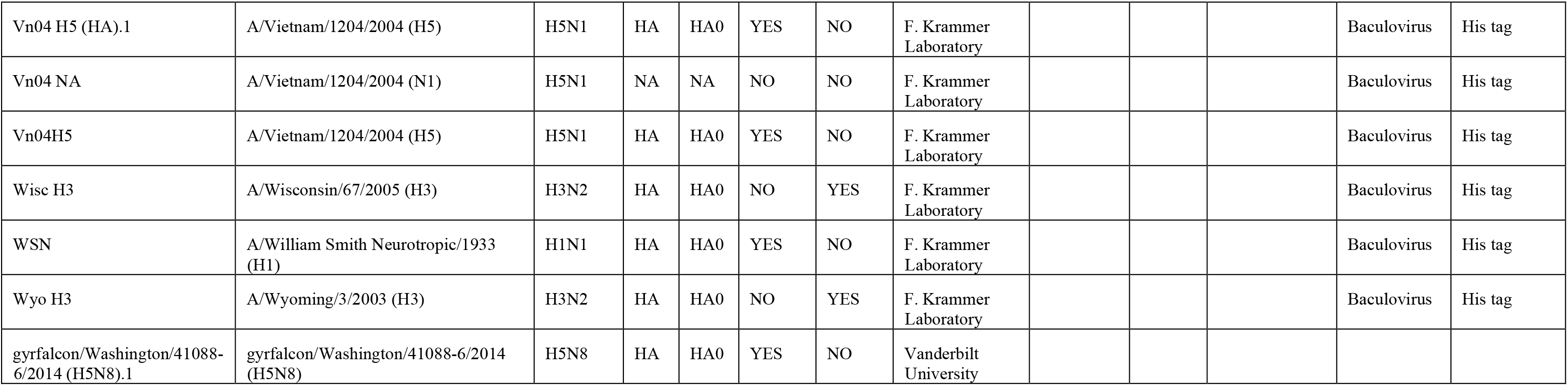
List of Proteins on Array #2.

